# SARS-CoV-2 genome sequencing from COVID-19 in Ecuadorian patients: a whole country analysis

**DOI:** 10.1101/2021.03.19.21253620

**Authors:** Sully Márquez, Belén Prado-Vivar, Juan José Guadalupe, Mónica Becerra-Wong, Bernardo Gutierrez, Clinical COVID-19 Ecuador Consortium, Juan Carlos Fernández-Cadena, Derly Andrade-Molina, Gabriel Morey-Leon, Miguel Moncayo, Rommel Guevara, Josefina Coloma, Gabriel Trueba, Michelle Grunauer, Verónica Barragán, Patricio Rojas-Silva, Paúl Cárdenas

## Abstract

SARS-CoV-2, the etiological agent of COVID-19, was first described in Wuhan, China in December 2019 and has now spread globally. Ecuador was the second country in South America to confirm cases and Guayaquil was one of the first cities in the world to experience high mortality due to COVID-19. The aim of this study was to describe the lineages circulating throughout the country and to compare the mutations in local variants, to the reference strain. In this work we used the MinION platform (Oxford Nanopore Technologies) to sequence the whole SARS-CoV-2 genomes of 119 patients from all provinces of Ecuador, using the ARTIC network protocols. Our data from lineage assignment of the one hundred and nineteen whole genomes revealed twenty different lineages. All genomes presented differences in the S gene compared to the Wuhan reference strain, being the D614G amino acid replacement the most common change. The B.1.1.119 lineage was the most frequent and was found in several locations in the Coast and Andean region. Three sequences were assigned to the new B.1.1.7 lineage. Our work is an important contribution to the understanding of the epidemiology of SARS-CoV-2 in Ecuador and South America.

**Highlights:**

- All 119 genomes showed mutations compared to the reference strain, which could be important to understand the virulence, severity and transmissibility of the virus.
- Until January 17, three sequences were assigned to the new B.1.1.7 lineage.
- Our findings suggest that there were at least twenty independent introductions of SARS-CoV-2 to Ecuador.

**Article Summary Line:** We report 119 sequences of SARS-CoV-2 across all Ecuadorian provinces, 20 different lineages were found until January 17^th^, including B.1.1.7.

## INTRODUCTION

SARS-CoV-2, the etiologic agent of COVID-19, has spread globally reaching all continents (1). Ecuador was the second country in South America, after Brazil, to report its first confirmed case, on February 29, 2020 (2). The number of cases has grown ever since, reaching 241,567 laboratory qPCR confirmed cases and 14,668 deaths by Jan 28, 2021 (3). The pandemic has had a severe impact in the Ecuadorian population. It is suggested that up to 838.35 people per million inhabitants may have died since the first COVID-19 case. Despite of this being the official data, mortality rate could be bias due to scarce testing.

Understanding the local epidemiology of SARS-CoV-2 can be greatly enhanced from understanding the viral evolution, establish the origin of variants (4), transmission of variants across different regions (5), the genetic diversity of variants in the population and to identify notable mutations (6). Additionally, key biological aspects such as virulence, transmissibility and infectivity of the circulating lineages can be investigated, when combined with clinical data from the patients (5). The importance of genomic surveillance is underscored with the current emergence of variants with potential increased transmission or disease severity like the B.1.1.7, P.1 and B.1.351. As a global effort, over 751,513 genome sequences of SARS-CoV-2 have been reported to date globally and are available in GenBank and the GISAID (Global Initiative on Sharing All Influenza Data) repositories (7).

Previous to this work there is no information about the SARS-CoV-2 lineages circulating in Ecuador. Here, we report one hundred and nineteen SARS-CoV-2 whole genome sequences sampled from every province of the country in the coastal, Andean and Amazonian regions plus the Galapagos archipielago. We identify lineages that circulated among the population from March 2020 through January 2021. We also describe the mutations present in these genomes as they could influence virulence, transmission and infectivity. The sequencing was performed using the portable MinION platform (Oxford Nanopore Technologies).

## MATERIALS AND METHODS

### Epidemiological information and sample collection

Nasopharyngeal swabs (NS) or broncho-alveolar lavage (BAL) samples were collected from patients in public third-level hospitals located in different provinces of Ecuador. Sample positivity for SARS-CoV-2 using standard RT-qPCR protocols, was officially reported to hospitals by the Ecuadorian Ministry of Public Health (MSP) and National Institute of Public Health and Research (INSPI). The use of samples was approved by the Bioethics Committee of Universidad San Francisco de Quito (CEISH No. 1234). The BAL samples were collected in a sterile tube with 2X DNA/RNA Shield (Zymo), and NS were immersed in 1X DNA/RNA Shield (Zymo), to ensure virus inactivation and preservation of the genetic material. Samples were transported immediately at 4°C to the Institute of Microbiology at USFQ (IM-USFQ) in a sealed container with all the biosecurity and containment measures recommended by the CDC of the USA (https://www.fda.gov/media/134922/download).

### RNA extraction

The genetic material from samples was extracted in a biosafety type II chamber with HEPA filters in the Virology Laboratory at IM-USFQ. The SV Total RNA Isolation System (Promega, USA) and Quick RNA viral, w/zymo-Spin IC (Zymo, USA) kits were used to extract RNA from samples for whole genome sequencing. A pre-digestion step was added to the RNA extraction protocol of the BAL samples as follows. Before nucleic acid extraction, 280 µl of the BAL samples were predigested with 360 µl of PureLink™ Genomic Lysis Buffer and 20 µl of proteinase K. The mix was incubated at 55°C for 10 minutes vortexing every 5 minutes (Life Technologies, USA). All RNA extractions were performed following manufacturer instructions. Retro-transcription was carried out using the Protocol of the Public Health England Genomics Lab (8,9) at the USFQ Bioinformatics Center and ARTIC protocol (10).

### Viral whole genome sequencing

The Primer Scheme (V1 and V3) developed by the ARTIC network for nCoV-2019 was employed to generate an amplicon tiling path across the viral genome (10,11). The final product of multiplex PCR was quantified using Qubit dsDNA HS (High Sensitivity) Assay Kit (Thermo Scientific, Invitrogen, Carlsbad, CA, USA). cDNA MinION library preparation was performed using the Rapid Barcoding kit (SQK-RBK004), native barcoding kit (NB-114) with ligation sequencing kit (LSK-109) following manufacturer instructions and then loaded into a MinION flow cell (FLO-MIN 106). Basecalling of FAST5 files was performed using Guppy (version 3.4.5) (12) (Oxford Nanopore Technologies). Also, the RAMPART software (v1.0.5) from the ARTIC Network (https://github.com/artic-network/rampart) was used to monitor sequencing in real-time. Sequence quality scoring, demultiplexing and adapter removal was performed with the NanoPlot (13) and Porechop (version 0.2.4) algorithms, respectively (https://github.com/rrwick/Porechop). The ARTIC Network bioinformatics pipeline was used for variant calling, and the reads were mapped against the reference strain Wuhan-Hu-1 (GenBank accession number MN908947), to generate consensus genomes. Tablet alignment viewer (version 1.19.09.3) (https://ics.hutton.ac.uk/tablet) was used to visualize the mapped sequence. The online tool NextClade (14) was used to determine the genomes clades. Then, genomes were uploaded to the CoV-GLUE resource (15), to determine the mutations, epidemiological linkage of circulating SARS-CoV-2 variants and primer mismatches. Lineage classification was carried out with Pangolin online software (https://github.com/cov-lineages/pangolin). Visualization of sequences in a phylogenetic tree was performed using the following strategy. The 119 Ecuadorian SARS COV 2 genomes of the present study were aligned with eight hundred and sixty one genomes from GISAID using NextAlign tool. A maximum likelihood phylogenetic reconstruction was performed with GTR substitution model and 1000 bootstrap resampling using IQtree 2.1.1. The phylogenetic tree was visualized with iTOL tool.

## RESULTS

### Complete genome sequencing revealed multiple entries of the virus to Ecuador

Sequencing results demonstrated that there were numerous SNPs in the ORF1a, ORF3a, ORF 7a, ORF7b, ORF8, S, M and N genes **(Figure 1 and Supplementary Table 1);** interestingly, the number of SNPs in the E gene was null. Our results confirmed the presence of several hotspot mutations in different viruses infecting the Ecuadorian communities. Phylogenetic analysis showed that Ecuadorian genomes were clustered with genomes from other countries, suggesting multiple entries **(Supplementary figure 1)**

**Figure 1.**
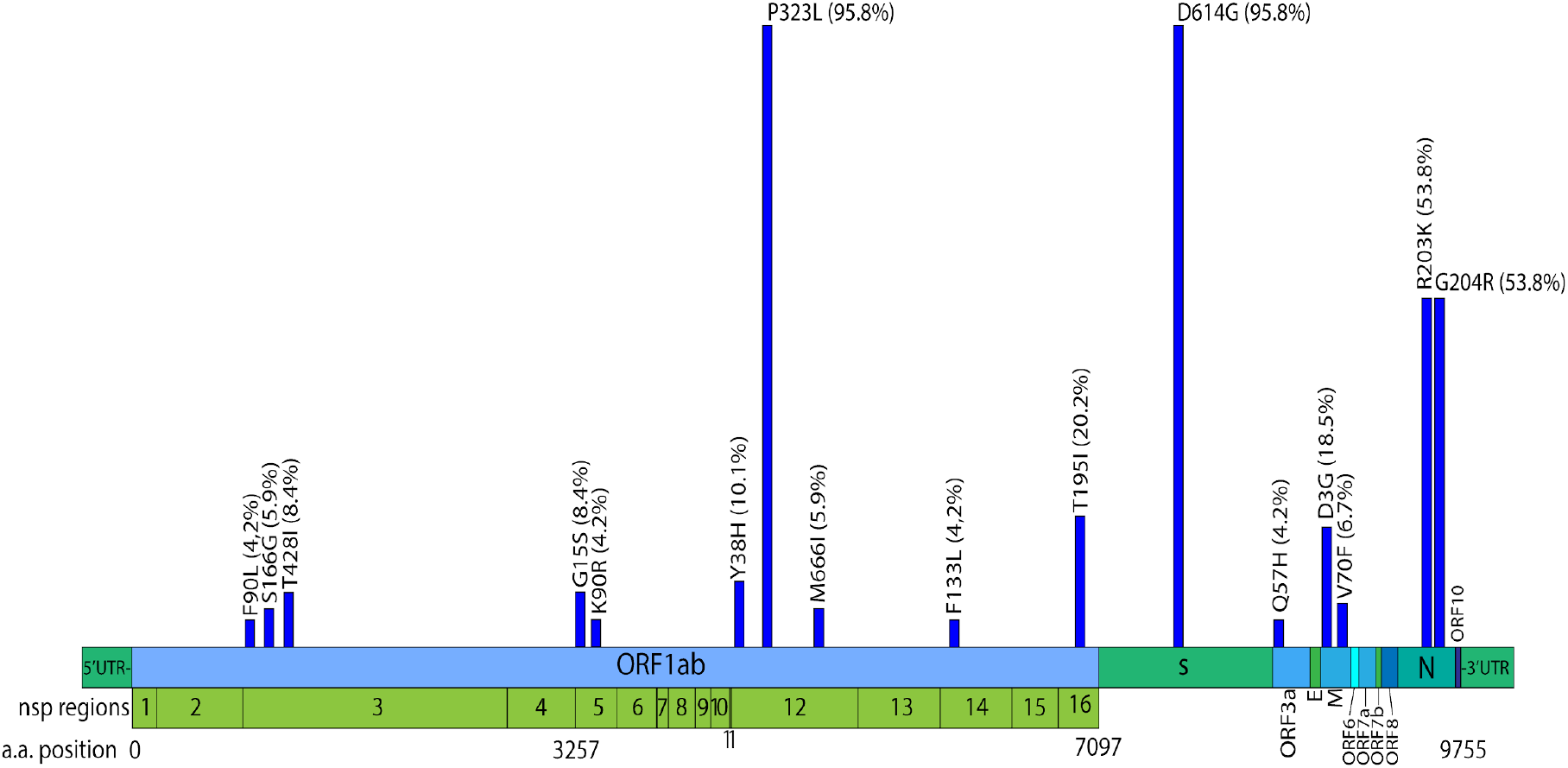
Genome annotation and frequency of the amino acid changes identified in at least 4% of the sequences. The amino acid changes are defined in comparison with the Wuhan-Hu-1 reference genome (GenBank accession number MN908947).

Twenty lineages were identified in which the dominant lineages were: B.1 (n=31) followed by B.1.1.119 (n=30), B.1.1.1 (n=10), B.1.1.207 (n=8), B.1.1.10 (n=6) and B.1.1.67 (n=5) (see Table S1 for all lineages). Interestingly some lineages were found only in 4 of the 24 provinces: A.1, B.1.9, B.1.371, B (Pichincha), A (Santo Domingo), B.1.308 (Zamora-Chinchipe) and B.1.6, B.1.325 (Imbabura). Lineages B.1.1.119 and B.1 were found in Awa and Waorani Amazonian indigenous communities respectively **(Table 1), (Supplementary Table 1)**.

**Table 1.** Lineages distribution in the 24 provinces of Ecuador.

| Province | Total population* | Total confirmed cases** | Cases per 100,00 hab | Lineages identified* |
| --- | --- | --- | --- | --- |
| Azuay | 881,394 | 15,383 | 1,745.30 | B.1.1.1, B.1.223, B.1 (n=4) |
| Bolívar | 209,933 | 2,866 | 1,365.20 | B.1.1.1, B.1.1.119 (n=3) |
| Cañar | 281,396 | 3,321 | 1,180.19 | B.1.1.119 (n=1) |
| Carchi | 186,869 | 4,725 | 2,528.51 | B.1.1.119 (n=1) |
| Cotopaxi | 488,716 | 4,076 | 777.86 | B.1, B.1.197 (n=2) |
| Chimborazo | 524,004 | 6,84 | 1,399.59 | B.1.1.207 (n=3) |
| Esmeraldas | 643,654 | 10,023 | 1,400.35 | B.1.223, B.1.1.119, B.1 (n=6) |
| El Oro | 715,751 | 5,361 | 832.90 | B.1.1.1, B.1 (n=4) |
| Galápagos | 33,042 | 1,211 | 3,665.03 | B.1 (n=1) |
| Guayas | 4,387,434 | 30,882 | 703.87 | B.1.1.1, B.1.1.119, B.1.1.10, B.1 (n=6) |
| Imbabura | 476,257 | 7,188 | 1,509.27 | B.1.1.207, B.1, B.1.67, B.1.1.119, B.1.223, B.1, B.1.1.1, B.1.6, B.1.1.31, B.1.1.10, B.1.325 (n=23) |
| Loja | 521,154 | 8,694 | 1,668.22 | B.1, B.1.1, B.1.1.119 (n=5) |
| Los Ríos | 921,763 | 5,713 | 619.79 | B.1.1.119, B.1.1.207, B.1, B.1.67, B.1.223, B.1.1.1, B.1.1.7 (n=14) |
| Manabí | 1,562,079 | 16,94 | 1,084.45 | B.1.1.119, B.1 (n=3) |
| Morona Santiago | 196,535 | 4,112 | 2,092.25 | B.1.1.119, B.1.1 (n=2) |
| Napo | 133,705 | 1,888 | 1,412.06 | B.1.1.119, B.1 (n=3) |
| Orellana | 161,338 | 2,167 | 1,343.14 | B.1.223, B.1 (n=4) |
| Pastaza | 114,202 | 2,501 | 2,189.98 | B.1.1.119 , B.1 (n=2) |
| Pichincha | 3,228,233 | 84,367 | 2,613.41 | B, B.1.1.119, B.1.14, B.1.371, B.1, |
|  |  |  |  | B.1.67, A.1, B.1.223, B.1.9, (n=14) |
| Santa Elena | 401,178 | 2,979 | 742.56 | B.1 (n=1) |
| Santo Domingo de los Tsáchilas | 458,58 | 6,61 | 1,441.41 | A, B.1.1.31, B.1.1.10, B.1.1.159 (n=7) |
| Sucumbíos | 230,503 | 3,183 | 1,380.89 | B.1, B.1.1.159 (n=3) |
| Tungurahua | 590,6 | 8,775 | 1,485.78 | B.1.1.119, B.1.197, B.1.1.10 (n=3) |
| Zamora Chinchipe | 120,416 | 1,762 | 1,463.26 | B.1, B.1.308, B.1.223 (n=4) |
| **Total** | **17,510,643** | **241,567** | **1,379.54** | **20 lineages (N=119)** |

\*Data based on the last estimations for 2020 from INEC (Instituto Nacional de
Estadísticas y Censos), \*\*Data from the official Ecuadorian portal
CoronavirusEcuador.com on January, 3 2021
https://www.coronavirusecuador.com/datos-provinciales/

Three of the analyzed sequences showed the aminoacid changes T183I, A890D, I1412T, P323L, L493F, N501Y, T553I, A570D, D614G, P681H, T716I, S982A, D1118H, Q27, R52I, Y73C, D3L, R203K, G204R and S235F placing them in the B.1.1.7 lineage. All sequences showed mutations at the S gene, being the most prevalent A23403G (95.79% sequences) conferring the D614G aminoacid change. **(Supplementary Table 1) (Table 2)**.

**Table 2.** Amino Acid replacement in gene S found in SARS-CoV2 genomes from Ecuador.

| Aminoacid replacement in S protein | no. of genomes with replacement (%) |
| --- | --- |
| A288S | 2 (1.68) |
| D614G | 114 (95.80) |
| E1207V | 1 (0.84) |
| L18F | 1 (0.84) |
| N1187S | 1 (0.84) |
| A570D | 3 (2.52) |
| D1118H | 3 (2.52) |
| G1167A | 4 (3.36) |
| N501Y | 3 (2.52) |
| Non-codon-aligned deletion | 3 (2.52) |
| P681H | 3 (2.52) |
| S982A | 3 (2.52) |
| T553I | 3 (2.52) |
| T716I | 3 (2.52) |

Primer mismatches were analyzed with seven COVID-19 diagnostic kits. All sequences showed changes at RdRP gene of Charité RT-PCR kit in positions 15469-15494, 15505-15530, and15431-15452.Mismatches were also found for the National Institute of Infectious Diseases Japan, CDC USA and CDC China RT-PCR diagnostic kits, **Supplementary Table 2**.

## DISCUSSION

Ecuador is one of the countries that has been severely affected by SARS-COV2 (1). Despite of the non-pharmaceutical interventions for preventing its dissemination, the virus continued to spread in all provinces. As most Latin American countries, Ecuador has had limited success in lowering transmission curves, mostly due to limited testing and socio-economic reasons(16). Public and private hospitals in Ecuador have continuously been flooded with patients with severe COVID-19 during the last year. In this study, we analyzed 119 SARS-CoV-2 whole genome sequences from all 24 provinces, using the third-generation sequencing MinION platform (Oxford Nanopore).

A total of twenty lineages were found in the 24 provinces. Unique lineages were detected only in Pichincha, Imbabura (Andes), Santo Domingo (Foothills) and Zamora-Chinchipe (Southern Amazon) provinces, however we cannot stablish that these lineages are not present in other provinces due to the limited genomic surveillance in the country Additionally, HGSQ-USFQ-007 and HGSQ-USFQ-010 genomes showed a distinct set of mutations. A mutation which may be increasing the virulence due to inhibition of the interferon response *in vitro* was found in these genomes (17)

SARS-COV2 is constantly changing through mutations, that led to the occurrence of new variants. Mutations in the S gene that confer a selective advantage to the virus had become more common. An example is A23403G mutation (D614G amino acid change) which was present in 95.79% of sequences from this study (18).

However, from all the variants that have been detected worldwide, only a small number are of public health concern. Three variants of concern (VOC) had emerged: B.1.1.7 in England, B.1.351 in South Africa, and P.1 in Brazil considering their improvements in viral replication mechanisms, transmissibility, and capacity to evade the immune host response. All these factors could have a direct impact on the efficacy of vaccines (18,19). In Ecuador B.1.1.7 was identified in three sequences from Los Ríos province, introduced by travelers from other countries Currently local authorities have reported that this variant is showing community-acquired transmission (20).

The presence of multiple lineages of SARS-COV-2 in Ecuador could be explained by independent introductions from different countries. The genomic surveillance results offer a high-resolution picture of the virus spread in the community probably due to the frequent movement of people between provinces. The current pandemic is unlikely to be the last, and it is, therefore, essential to improve the response capacity of our public health systems and to implement and strengthen continuous scientific research programs. Only in this way, we will be able to better understand these types of threats, act based on evidence, and thus reduce their impact. Community-acquired transmission is widespread in Ecuador and more samples from community-acquired infections are needed to inform analyses of the local epidemiology of the virus.

Medical and scientific efforts throughout the country made possible to compile at least one sequence from each province, however we must join efforts with other universities and state laboratories to increase genomic surveillance in the country

We are aware that the small number of samples reported in this study might not capture the full picture of what is happening in Ecuador. However, revealing the information contained in the SARS-CoV-2 virus genome sequences is a robust tool to understand the epidemiology of COVID-19 locally. The present study is a steppingstone to understand the importance of carrying local genomic surveillance for infectious diseases circulating in the country.

## Data Availability

All data are freely available on GISAID.org.

## ACKNOWLEDGEMENTS

This work was funded by Universidad San Francisco de Quito through Emergency Research Funds and COCIBA grants (P.R-S.) and CADDE project (www.caddecentre.org/). P.C. is funded by NIH FIC D43TW010540 Global Health Equity Scholars. Lourdes Torres, Diego Quiroga and the people of USFQ-Proyectos for their support on the project development.

We acknowledge the submitting authors and institutions of the sequences of SARS-CoV-2 from GISAID Database.

## CONSORTIA

The members of Clinical COVID-19 Ecuador Consortium in alphabetical order are Alejandra Ramones, Alexandra Tino, Andrea Carrera, Andrea Macias, Anita Garcia, Carlos Guerrero, Carlos Mena, Carlos Tobar, Carolina Proaño-Bolaños, Damaris Zandoya, Dayron Brossard, Diana Zambrano, Diego Egas, Eddy Chavez, Edison Chavez, Edison Ligña, Edy Quizhpe, Eulalia Pazmiño, Fabian Aguilar, Fausto Maldonado, Francisco Córdova, Francisco Mora, Francisco Rodriguez, Franklin Espinoza, Freddy Iza, Fredy Loor, Gabriel Morey, Geovanny Cazorla, Giovanna Moran, Grace Salazar, Hermelinda Paguay, Jonathan Araujo, Jorge Luis Velez, Jorge Montaño, Jorge Reyes, Juan Gaviria, Juan Zuñiga, Karina Barragan, Karolina Pacheco, Katherine Apunte, Khurram Mahbbob, Killen Briones-Claudette, Killen Briones-Zamora, Ligia Briceño, Manuel Jaramillo, Manuel Jibaja, Marcelo Ortiz, Marcos Di Stefano, Maureen Mosquera, Milton Tobar, Nabih Dahik, Nina Espinoza de los Monteros, Ninfa Henriquez, Patricio Reyes, Rene Bracho, Rosario Erazo, Sonia Sislema, Stalin Castillo, Stephanie Arregui, Tania Guayasamin, Yeimy Rojas and Yomara Napa.

## AUTHOR CONTRIBUTIONS

Conceptualization: M.J., G.T., M.G., P.C. Methodology: S.M., B.P.-V., J.J.G., M.T., P.C. Software: B.P.-V., V.B., M.B., P.C. Validation: B.P.-V., V.B. and P.C. Formal analysis and investigation: V.B., P.R.-S., M.B, P.C. Writing - Original Draft: P.R.-S. and P.C. Writing - Review & Editing: All authors. Visualization: V.B. and P.C. and M.A Supervision: G.T. Funding acquisition: M.G, G.T.,J.C & P.C. The Clinical COVID-19 Ecuador Consortium collected samples and provided epidemiological information for all the sequences.

## Ethical approval

Ethical approval for all samples was given by CEISH-USFQ (Comité de Ética de Investigación en Seres Humanos-USFQ): IE-JP067-2020-CEISH-USFQ. Additionally by the Ministerio de Salud Publica Comité Expedito (021-2020)

## Informed consent

Every patient or relative (in case of severely ill patients) has signed the informed consent.

## Biographical Sketch

SM is a graduate student at the Instituto de Microbiología in Universidad San Francisco de Quito and interested in virology research.

## Figures and tables legends

**Supplementary table 1**. Mutations found in four SARS CoV2 Genomes from Ecuador compared to Wuhan-Hu-1 (GenBank accession number MN908947)

**Supplementary table 2**. Primer mismatches analyzed with seven COVID 19 diagnostic kits.

**Supplementary figure 1.**
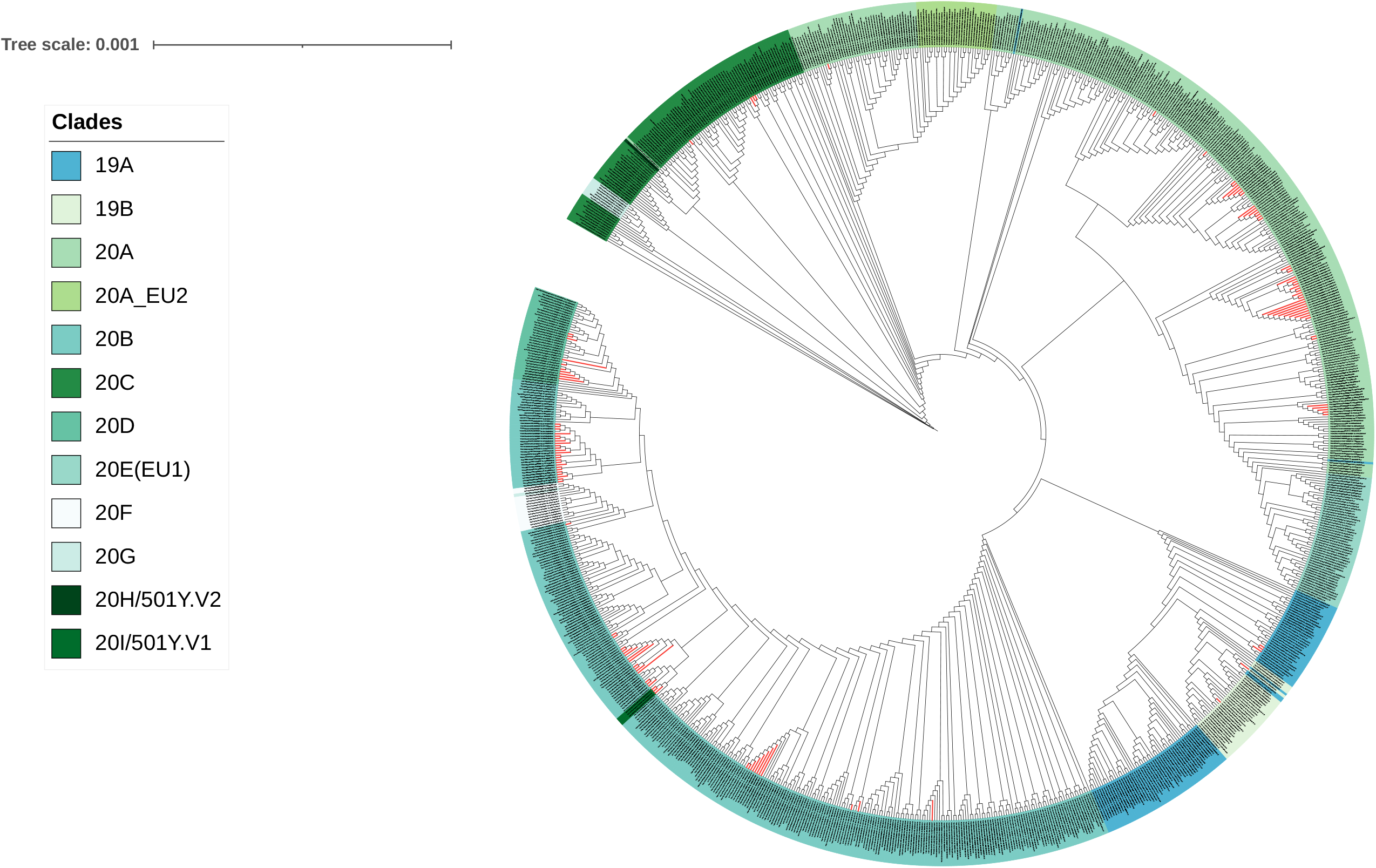
Phylogenetic analysis of SARS-COV2 genomes. The phylogenetic tree was generated using the software IQ tree 2.1.1 and was visualized and annotated using the iTOL tool. Ecuador sequences are highlighted in red.

