## Supplemental Table 2 for "SARS-CoV-2 genome sequencing from COVID-19 in Ecuadorian patients: a whole country analysis"

**Supplementary table 2.** Primer mismatches analyzed with seven COVID 19 diagnostic kits.

| Sequence ID | GISAID ID | China CDC RT-PCR | | HKU RT-PCR | | Charité RT-PCR | | Ministry of Public Health Thailand RT-PCR | National Institute of Infectious Diseases Japan Rt-PCR | USA CDC RT-PCR | | | Rt- PCR N gene 2020 |
|---|---|---|---|---|---|---|---|---|---|---|---|---|---|
| | | N | ORF1ab | N | ORF1b-nsp14 | E | RdRP | N | N | N1 | N2 | N3 | N |
| USFQ-185 | EPI_ISL_697784 | - | - | - | - | - | Genome position:15469-15494 Mismatches: 2, R15480C*, T15489A* Genome position:15505-15530 Mismatches: 1, S15519T* | - | Genome position:29263-29282 Mismatches: 1, C29277G* | - | - | - | - |
| USFQ-1188 | EPI_ISL_697785 | - | - | - | - | - | Genome position:15469-15494 Mismatches: 2, R15480C*, T15489A* | - | Genome position:29263-29282 Mismatches: 1, C29277G* | - | - | - | - |
| USFQ-249 | EPI_ISL_697787 | - | - | - | - | - | Genome position:15469-15494 Mismatches: 2, R15480C*, T15489A* | - | Genome position:29263-29282 Mismatches: 1, C29277G* | - | - | - | - |
| USFQ-322 | EPI_ISL_697788 | - | - | - | - | - | Genome position:15469-15494 Mismatches: 2, R15480C*, T15489A* | - | Genome position:29263-29282 Mismatches: 1, C29277G* | - | - | - | - |
| USFQ-555 | EPI_ISL_728203 | - | - | - | - | - | Genome position:15469-15494 Mismatches: 2, R15480C*, T15489A* | - | Genome position:29263-29282 Mismatches: 1, C29277G* | Genome position:28287-28306 Mismatches: 1, C28290T | - | - | - |
| USFQ-556 | EPI_ISL_728204 | - | - | - | - | - | Genome position:15469-15494 Mismatches: 2, R15480C*, T15489A* | - | Genome position:29263-29282 Mismatches: 1, C29277G* | - | - | - | - |
| USFQ-558 | EPI_ISL_728205 | - | - | - | - | - | Genome position:15469-15494 Mismatches: 2, R15480C*, T15489A* | - | Genome position:29263-29282 Mismatches: 1, C29277G* | Genome position:28287-28306 Mismatches: 1, C28290T | - | - | - |
| USFQ-220 | EPI_ISL_539790 | - | - | - | - | - | Genome position:15469-15494 Mismatches: 2, R15480C*, T15489A* | - | Genome position:29263-29282 Mismatches: 1, C29277G* | - | - | - | - |
| USFQ-222 | EPI_ISL_539791 | - | - | - | - | - | Genome position:15469-15494 Mismatches: 2, R15480C*, T15489A* | - | Genome position:29263-29282 Mismatches: 1, C29277G* | - | - | - | - |
| USFQ-224 | EPI_ISL_539792 | - | - | - | - | - | Genome position:15469-15494 Mismatches: 2, R15480C*, T15489A* | - | Genome position:29263-29282 Mismatches: 1, C29277G* | - | - | - | - |
| USFQ-226 | EPI_ISL_539793 | - | - | - | - | - | Genome position:15469-15494 Mismatches: 2, R15480C*, T15489A* | - | Genome position:29263-29282 Mismatches: 1, C29277G* | - | - | - | - |
| USFQ-253 | EPI_ISL_574431 | - | - | - | - | - | Genome position:15469-15494 Mismatches: 2, R15480C*, T15489A* | - | Genome position:29263-29282 Mismatches: 1, C29277G* | - | - | - | - |
| USFQ-1026 | EPI_ISL_594118 | - | - | - | - | - | Genome position:15469-15494 Mismatches: 2, R15480C*, T15489A* | - | Genome position:29263-29282 Mismatches: 1, C29277G* | - | - | - | - |
| USFQ-228 | EPI_ISL_660529 | - | - | - | - | - | Genome position:15469-15494 Mismatches: 2, R15480C*, T15489A* | - | Genome position:29263-29282 Mismatches: 1, C29277G* | - | - | - | - |
| USFQ-230 | EPI_ISL_660530 | - | - | - | - | - | Genome position:15469-15494 Mismatches: 2, R15480C*, T15489A* | - | Genome position:29263-29282 Mismatches: 1, C29277G* | - | - | - | - |
| USFQ-231 | EPI_ISL_660531 | - | - | - | - | - | Genome position:15469-15494 Mismatches: 2, R15480C*, T15489A* | - | Genome position:29263-29282 Mismatches: 1, C29277G* | - | - | - | - |
| USFQ-402 | EPI_ISL_660532 | - | - | - | - | - | Genome position:15469-15494 Mismatches: 2, R15480C*, T15489A* | - | Genome position:29263-29282 Mismatches: 1, C29277G* | - | - | - | - |
| USFQ-485 | EPI_ISL_660533 | - | - | - | - | - | Genome position:15469-15494 Mismatches: 2, R15480C*, T15489A* | - | Genome position:29263-29282 Mismatches: 1, C29277G* | - | - | - | - |
| USFQ-487 | EPI_ISL_660534 | - | - | - | - | - | Genome position:15469-15494 Mismatches: 2, R15480C*, T15489A* | - | Genome position:29263-29282 Mismatches: 1, C29277G* | - | - | - | - |
| USFQ-506 | EPI_ISL_660535 | - | - | - | - | - | Genome position:15469-15494 Mismatches: 2, R15480C*, T15489A* | - | Genome position:29263-29282 Mismatches: 1, C29277G* | - | - | - | - |
| USFQ-509 | EPI_ISL_660536 | - | - | - | - | - | Genome position:15469-15494 Mismatches: 2, R15480C*, T15489A* | - | Genome position:29263-29282 Mismatches: 1, C29277G* | - | - | - | - |
| USFQ-513 | EPI_ISL_660537 | - | - | - | - | - | Genome position:15469-15494 Mismatches: 2, R15480C*, T15489A* | - | Genome position:29263-29282 Mismatches: 1, C29277G* | - | - | - | - |
| USFQ-515 | EPI_ISL_660538 | - | - | - | - | - | Genome position:15469-15494 Mismatches: 2, R15480C*, T15489A* | - | Genome position:29263-29282 Mismatches: 1, C29277G* | - | - | - | - |
| USFQ-520 | EPI_ISL_660539 | - | - | - | - | - | Genome position:15469-15494 Mismatches: 2, R15480C*, T15489A* | - | Genome position:29263-29282 Mismatches: 1, C29277G* | - | - | - | - |
| USFQ-189 | EPI_ISL_697786 | - | - | - | - | - | Genome position:15469-15494 Mismatches: 2, R15480C*, T15489A* | - | Genome position:29263-29282 Mismatches: 1, C29277G* | - | - | - | - |
| USFQ-346 | EPI_ISL_697790 | - | - | - | - | - | Genome position:15469-15494 Mismatches: 2, R15480C*, T15489A* | - | Genome position:29263-29282 Mismatches: 1, C29277G* | - | - | - | - |
| USFQ-405 | EPI_ISL_697792 | - | - | - | - | - | Genome position:15469-15494 Mismatches: 2, R15480C*, T15489A* | - | Genome position:29263-29282 Mismatches: 1, C29277G* | - | - | - | - |
| USFQ-433 | EPI_ISL_697793 | - | - | - | - | - | Genome position:15469-15494 Mismatches: 2, R15480C*, T15489A* | - | Genome position:29263-29282 Mismatches: 1, C29277G* | - | - | - | - |
| USFQ-524 | EPI_ISL_697795 | - | - | - | - | - | Genome position:15469-15494 Mismatches: 2, R15480C*, T15489A* | - | Genome position:29263-29282 Mismatches: 1, C29277G* | - | - | - | - |
| USFQ-527 | EPI_ISL_697796 | - | - | - | - | - | Genome position:15469-15494 Mismatches: 2, R15480C*, T15489A* | - | Genome position:29263-29282 Mismatches: 1, C29277G* | - | - | - | - |
| USFQ-528 | EPI_ISL_697797 | - | - | - | - | - | Genome position:15469-15494 Mismatches: 2, R15480C*, T15489A* | - | Genome position:29263-29282 Mismatches: 1, C29277G* | - | - | - | - |
| USFQ-531 | EPI_ISL_697798 | - | - | - | - | - | Genome position:15469-15494 Mismatches: 2, R15480C*, T15489A* | - | Genome position:29263-29282 Mismatches: 1, C29277G* | - | - | - | - |
| USFQ-533 | EPI_ISL_697799 | - | - | - | - | - | Genome position:15469-15494 Mismatches: 2, R15480C*, T15489A* | - | Genome position:29263-29282 Mismatches: 1, C29277G* | - | - | - | - |
| USFQ-539 | EPI_ISL_697800 | - | - | - | - | - | Genome position:15469-15494 Mismatches: 2, R15480C*, T15489A* | - | Genome position:29263-29282 Mismatches: 1, C29277G* | - | - | - | - |
| USFQ-553 | EPI_ISL_728202 | - | - | - | - | - | Genome position:15469-15494 Mismatches: 2, R15480C*, T15489A* | - | Genome position:29263-29282 Mismatches: 1, C29277G* | Genome position:28287-28306 Mismatches: 1, C28290T | - | - | - |
| USFQ-581 | EPI_ISL_803098 | - | - | - | - | - | Genome position:15469-15494 Mismatches: 2, R15480C*, T15489A* | - | Genome position:29263-29282 Mismatches: 1, C29277G* | - | - | - | - |
| USFQMC01 | EPI_ISL_525438 | - | - | - | - | - | Genome position:15469-15494 Mismatches: 2, R15480C*, T15489A* Genome position:15431-15452 Mismatches: 1, G15438T Genome position:15505-15530 Mismatches: 1, S15519T* | - | Genome position:29263-29282 Mismatches: 1, C29277G* | - | - | - | - |
| USFQ-193 | EPI_ISL_527809 | - | - | - | - | - | Genome position:15469-15494 Mismatches: 2, R15480C*, T15489A* | - | Genome position:29263-29282 Mismatches: 1, C29277G* | - | - | - | - |
| USFQ-194 | EPI_ISL_527810 | - | - | - | - | - | Genome position:15469-15494 Mismatches: 2, R15480C*, T15489A* | - | Genome position:29263-29282 Mismatches: 1, C29277G* | - | - | - | - |
| USFQ-195 | EPI_ISL_527811 | - | - | - | - | - | Genome position:15469-15494 Mismatches: 2, R15480C*, T15489A* | - | Genome position:29263-29282 Mismatches: 1, C29277G* | - | - | - | - |
| USFQ-196 | EPI_ISL_527812 | - | - | - | - | - | Genome position:15469-15494 Mismatches: 2, R15480C*, T15489A* | - | Genome position:29263-29282 Mismatches: 1, C29277G* | - | - | - | - |
| USFQ-197 | EPI_ISL_527813 | - | - | - | - | - | "Genome position:15469-15494 Mismatches: 2, R15480C*, T15489A* | - | Genome position:29263-29282 Mismatches: 1, C29277G* | - | - | - | - |
| USFQ-199 | EPI_ISL_527814 | - | - | - | - | - | Genome position:15469-15494 Mismatches: 2, R15480C*, T15489A* | - | Genome position:29263-29282 Mismatches: 1, C29277G* | - | - | - | - |
| USFQ-201 | EPI_ISL_527815 | - | - | - | - | - | Genome position:15469-15494 Mismatches: 2, R15480C*, T15489A* | - | Genome position:29263-29282 Mismatches: 1, C29277G* | - | - | - | - |
| USFQ-203 | EPI_ISL_527816 | - | - | - | - | - | Genome position:15469-15494 Mismatches: 2, R15480C*, T15489A* | - | Genome position:29263-29282 Mismatches: 1, C29277G* | - | - | - | - |
| USFQ-204 | EPI_ISL_527817 | - | - | - | - | - | Genome position:15469-15494 Mismatches: 2, R15480C*, T15489A* | - | Genome position:29263-29282 Mismatches: 1, C29277G* | - | - | - | - |
| USFQ-206 | EPI_ISL_527818 | - | - | - | - | - | Genome position:15469-15494 Mismatches: 2, R15480C*, T15489A* | - | Genome position:29263-29282 Mismatches: 1, C29277G* | - | - | - | - |
| USFQ-207 | EPI_ISL_527819 | - | - | - | - | - | Genome position:15469-15494 Mismatches: 2, R15480C*, T15489A* | - | Genome position:29263-29282 Mismatches: 1, C29277G* | - | - | - | - |
| USFQ-208 | EPI_ISL_539783 | - | - | - | - | - | Genome position:15469-15494 Mismatches: 2, R15480C*, T15489A* | - | Genome position:29263-29282 Mismatches: 1, C29277G* | - | - | - | - |
| USFQ-209 | EPI_ISL_539784 | - | - | - | - | - | Genome position:15469-15494 Mismatches: 2, R15480C*, T15489A* | - | Genome position:29263-29282 Mismatches: 1, C29277G* | - | - | - | - |
| USFQ-212 | EPI_ISL_539785 | - | - | - | - | - | Genome position:15469-15494 Mismatches: 2, R15480C*, T15489A* | - | Genome position:29263-29282 Mismatches: 1, C29277G* | - | - | - | - |
| USFQ-213 | EPI_ISL_539786 | - | - | - | - | - | Genome position:15469-15494 Mismatches: 2, R15480C*, T15489A* | - | Genome position:29263-29282 Mismatches: 1, C29277G* | - | - | - | - |
| USFQ-214 | EPI_ISL_539787 | - | - | - | - | - | Genome position:15469-15494 Mismatches: 2, R15480C*, T15489A* | - | Genome position:29263-29282 Mismatches: 1, C29277G* | - | - | - | - |
| USFQ-216 | EPI_ISL_539788 | - | - | - | - | - | Genome position:15469-15494 Mismatches: 2, R15480C*, T15489A* | - | Genome position:29263-29282 Mismatches: 1, C29277G* | - | - | - | - |
| USFQ-219 | EPI_ISL_539789 | | | | | | Genome position:15469-15494 Mismatches: 2, R15480C*, T15489A* | - | Genome position:29263-29282 Mismatches: 1, C29277G* | - | - | - | - |
| USFQ-344 | EPI_ISL_697789 | | | | | | Genome position:15469-15494 Mismatches: 2, R15480C*, T15489A* | - | Genome position:29263-29282 Mismatches: 1, C29277G* | - | - | - | - |
| USFQ-381 | EPI_ISL_697791 | | | | | | Genome position:15469-15494 Mismatches: 2, R15480C*, T15489A* | - | Genome position:29263-29282 Mismatches: 1, C29277G* | - | - | - | - |
| USFQ-523 | EPI_ISL_697794 | | | | | | Genome position:15469-15494 Mismatches: 2, R15480C*, T15489A* | - | Genome position:29263-29282 Mismatches: 1, C29277G* | - | - | - | - |
| HEE-01 | EPI_ISL_417482 | - | - | - | - | - | Genome position: 15469-15494 Mismatches: 2, R15480C*, T15489A* | - | Genome position: 29263-29282 Mismatches: 1, C29277G* | - | - | - | - |
| HGSQ-USFQ-018 | EPI_ISL_422563 | - | - | - | - | - | Genome position: 15469-15494 Mismatches: 2, R15480C*, T15489A* | - | Genome position: 29263-29282 Mismatches: 1, C29277G* | - | - | - | - |
| HGSQ-USFQ-007 | EPI_ISL_422564 | - | - | - | - | - | Genome position: 15469-15494 Mismatches: 2, R15480C*, T15489A* | - | Genome position: 29263-29282 Mismatches: 1, C29277G* | - | - | - | - |
| HGSQ-USFQ-010 | EPI_ISL_422565 | - | - | - | - | - | Genome position: 15469-15494 Mismatches: 2, R15480C*, T15489A* | - | Genome position: 29263-29282 Mismatches: 1, C29277G* | - | - | - | - |
| USFQ-020 | EPI_ISL_471267 | - | - | - | - | - | Genome position: 15469-15494 Mismatches: 2, R15480C*, T15489A* | - | Genome position: 29263-29282 Mismatches: 1, C29277G* | - | - | - | - |
| USFQ-045 | EPI_ISL_471268 | - | - | - | - | - | Genome position: 15469-15494 Mismatches: 2, R15480C*, T15489A* | - | Genome position: 29263-29282 Mismatches: 1, C29277G* | - | - | - | - |
| USFQ-707 | EPI_ISL_471269 | - | - | - | - | - | Genome position: 15469-15494 Mismatches: 2, R15480C*, T15489A* | - | Genome position: 29263-29282 Mismatches: 1, C29277G* | - | - | - | - |
| USFQ-112 | EPI_ISL_471270 | - | - | - | - | - | Genome position: 15469-15494 Mismatches: 2, R15480C*, T15489A* | - | Genome position: 29263-29282 Mismatches: 1, C29277G* | - | - | - | - |
| USFQ-106 | EPI_ISL_471271 | - | - | - | - | - | Genome position: 15469-15494 Mismatches: 2, R15480C*, T15489A* | - | Genome position: 29263-29282 Mismatches: 1, C29277G* | - | - | - | - |
| USFQ-004 | EPI_ISL_477014 | - | - | - | - | - | Genome position: 15469-15494 Mismatches: 2, R15480C*, T15489A* | - | Genome position: 29263-29282 Mismatches: 1, C29277G* | - | - | - | - |
| USFQ-096 | EPI_ISL_477015 | - | - | - | - | - | Genome position: 15469-15494 Mismatches: 2, R15480C*, T15489A* | - | Genome position: 29263-29282 Mismatches: 1, C29277G* | - | - | - | - |
| USFQ-097 | EPI_ISL_477016 | - | - | - | - | - | Genome position: 15469-15494 Mismatches: 2, R15480C*, T15489A* | - | Genome position: 29263-29282 Mismatches: 1, C29277G* | - | - | - | - |
| USFQ-026 | EPI_ISL_481244 | - | - | - | - | - | Genome position: 15469-15494 Mismatches: 2, R15480C*, T15489A* | - | Genome position: 29263-29282 Mismatches: 1, C29277G* | - | - | - | - |
| USFQ-027 | EPI_ISL_481245 | - | - | - | - | - | Genome position: 15469-15494 Mismatches: 2, R15480C*, T15489A* | - | Genome position: 29263-29282 Mismatches: 1, C29277G* | - | - | - | - |
| USFQ-054 | EPI_ISL_481246 | - | - | - | - | - | Genome position: 15469-15494 Mismatches: 2, R15480C*, T15489A* | - | Genome position: 29263-29282 Mismatches: 1, C29277G* | - | - | - | - |
| USFQ-065 | EPI_ISL_481247 | - | - | - | - | - | Genome position: 15469-15494 Mismatches: 2, R15480C*, T15489A* | - | Genome position: 29263-29282 Mismatches: 1, C29277G* | - | - | - | - |
| USFQ-066 | EPI_ISL_481248 | - | - | - | - | - | Genome position: 15469-15494 Mismatches: 2, R15480C*, T15489A* | - | Genome position: 29263-29282 Mismatches: 1, C29277G* | - | - | - | - |
| USFQ-105 | EPI_ISL_486842 | - | - | - | - | - | Genome position: 15469-15494 Mismatches: 2, R15480C*, T15489A* | - | Genome position: 29263-29282 Mismatches: 1, C29277G* | - | - | - | - |
| USFQ-108 | EPI_ISL_486843 | - | - | - | - | - | Genome position: 15469-15494 Mismatches: 2, R15480C*, T15489A* | - | Genome position: 29263-29282 Mismatches: 1, C29277G* | - | - | - | - |
| USFQ-109 | EPI_ISL_486844 | - | - | - | - | - | Genome position: 15469-15494 Mismatches: 2, R15480C*, T15489A* | - | Genome position: 29263-29282 Mismatches: 1, C29277G* | - | - | - | - |
| USFQ-110 | EPI_ISL_486845 | - | - | - | - | - | Genome position: 15469-15494 Mismatches: 2, R15480C*, T15489A* | - | Genome position: 29263-29282 Mismatches: 1, C29277G* | - | - | - | - |
| USFQ-111 | EPI_ISL_486846 | - | - | - | - | - | Genome position: 15469-15494 Mismatches: 2, R15480C*, T15489A* | - | Genome position: 29263-29282 Mismatches: 1, C29277G* | - | - | - | - |
| USFQ-1112 | EPI_ISL_486847 | - | - | - | - | - | Genome position: 15469-15494 Mismatches: 2, R15480C*, T15489A* | - | Genome position: 29263-29282 Mismatches: 1, C29277G* | - | - | - | - |
| USFQ-113 | EPI_ISL_486848 | - | - | - | - | - | Genome position: 15469-15494 Mismatches: 2, R15480C*, T15489A* | - | Genome position: 29263-29282 Mismatches: 1, C29277G* | - | - | - | - |
| USFQ-114 | EPI_ISL_486849 | - | - | - | - | - | Genome position: 15469-15494 Mismatches: 2, R15480C*, T15489A* | - | Genome position: 29263-29282 Mismatches: 1, C29277G* | - | - | - | - |
| USFQ-118/ | EPI_ISL_486850 | - | - | - | - | - | Genome position: 15469-15494 Mismatches: 2, R15480C*, T15489A* | - | Genome position: 29263-29282 Mismatches: 1, C29277G* | - | - | - | - |
| USFQ-119 | EPI_ISL_486851 | - | - | - | - | - | Genome position: 15469-15494 Mismatches: 2, R15480C*, T15489A* | - | Genome position: 29263-29282 Mismatches: 1, C29277G* | - | - | - | - |
| USFQ-120 | EPI_ISL_491932 | - | - | - | - | - | Genome position: 15469-15494 Mismatches: 2, R15480C*, T15489A* | - | Genome position: 29263-29282 Mismatches: 1, C29277G* | - | - | - | - |
| USFQ-128 | EPI_ISL_491933 | - | - | - | - | - | Genome position: 15469-15494 Mismatches: 2, R15480C*, T15489A* | - | Genome position: 29263-29282 Mismatches: 1, C29277G* | - | - | - | - |
| USFQ-129 | EPI_ISL_491934 | - | - | - | - | - | Genome position: 15469-15494 Mismatches: 2, R15480C*, T15489A* | - | Genome position: 29263-29282 Mismatches: 1, C29277G* | - | - | - | - |
| USFQ-131 | EPI_ISL_491935 | - | - | - | - | - | Genome position: 15469-15494 Mismatches: 2, R15480C*, T15489A* | - | Genome position: 29263-29282 Mismatches: 1, C29277G* | - | - | - | - |
| USFQ-139 | EPI_ISL_491936 | - | - | - | - | - | Genome position: 15469-15494 Mismatches: 2, R15480C*, T15489A* | - | Genome position: 29263-29282 Mismatches: 1, C29277G* | - | - | - | - |
| USFQ-147 | EPI_ISL_491937 | - | - | - | - | - | Genome position: 15469-15494 Mismatches: 2, R15480C*, T15489A* | - | Genome position: 29263-29282 Mismatches: 1, C29277G* | - | - | - | - |
| USFQ-150 | EPI_ISL_491938 | - | - | - | - | - | Genome position: 15469-15494 Mismatches: 2, R15480C*, T15489A* | - | Genome position: 29263-29282 Mismatches: 1, C29277G* | - | - | - | - |
| USFQ-126 | EPI_ISL_491939 | - | - | - | - | - | Genome position: 15469-15494 Mismatches: 2, R15480C*, T15489A* | - | Genome position: 29263-29282 Mismatches: 1, C29277G* | - | - | - | - |
| USFQ-127 | EPI_ISL_491940 | - | - | - | - | - | Genome position: 15469-15494 Mismatches: 2, R15480C*, T15489A* | - | Genome position: 29263-29282 Mismatches: 1, C29277G* | - | - | - | - |
| USFQ-133 | EPI_ISL_491941 | - | - | - | - | - | Genome position: 15469-15494 Mismatches: 2, R15480C*, T15489A* | - | Genome position: 29263-29282 Mismatches: 1, C29277G* | - | - | - | - |
| USFQ-161 | EPI_ISL_516648 | Clade* | - | - | - | - | Genome position: 15469-15494 Mismatches: 2, R15480C*, T15489A* | - | Genome position: 29263-29282 Mismatches: 1, C29277G* | - | - | - | - |
| USFQ-162 | EPI_ISL_516649 | - | - | - | - | - | Genome position: 15431-15452 Mismatches:1, G15438T | - | Genome position: 29263-29282 Mismatches: 1, C29277G* | - | - | - | - |
| USFQ-165 | EPI_ISL_516650 | - | - | - | - | - | Genome position: 15469-15494 Mismatches: 2, R15480C*, T15489A* | - | Genome position: 29263-29282 Mismatches: 1, C29277G* | - | - | - | - |
| USFQ-167 | EPI_ISL_516651 | - | - | - | - | - | Genome position: 15431-15452 Mismatches:1, G15438T | - | Genome position: 29263-29282 Mismatches: 1, C29277G* | - | - | - | - |
| USFQ-171 | EPI_ISL_516652 | - | - | - | - | - | Genome position: 15431-15452 Mismatches:1, G15438T | - | Genome position: 29263-29282 Mismatches: 1, C29277G* | - | - | - | - |
| USFQ-163 | EPI_ISL_525430 | - | - | - | - | - | Genome position: 15469-15494 Mismatches: 2, R15480C*, T15489A* | - | Genome position: 29263-29282 Mismatches: 1, C29277G* | - | - | - | - |
| USFQ-177 | EPI_ISL_525431 | - | - | - | - | - | Genome position: 15469-15494 Mismatches: 2, R15480C*, T15489A* | - | Genome position: 29263-29282 Mismatches: 1, C29277G* | - | - | - | - |
| USFQ-178 | EPI_ISL_525432 | - | - | - | - | - | Genome position: 15469-15494 Mismatches: 2, R15480C*, T15489A* | - | Genome position: 29263-29282 Mismatches: 1, C29277G* | - | - | - | - |
| USFQ-182 | EPI_ISL_525433 | - | - | - | - | - | Genome position: 15469-15494 Mismatches: 2, R15480C*, T15489A* | - | Genome position: 29263-29282 Mismatches: 1, C29277G* | - | - | - | - |
| USFQ-183 | EPI_ISL_525434 | - | - | - | - | - | Genome position: 15469-15494 Mismatches: 2, R15480C*, T15489A* | - | Genome position: 29263-29282 Mismatches: 1, C29277G* | - | - | - | - |
| USFQ-184 | EPI_ISL_525435 | - | - | - | - | - | Genome position: 15469-15494 Mismatches: 2, R15480C*, T15489A* | - | Genome position: 29263-29282 Mismatches: 1, C29277G* | - | - | - | - |
| USFQ-186 | EPI_ISL_525436 | - | - | - | - | - | Genome position: 15431-15452 Mismatches:1, G15438T | - | Genome position: 29263-29282 Mismatches: 1, C29277G* | - | - | - | - |
| USFQ-188 | EPI_ISL_525437 | - | - | - | - | - | Genome position: 15469-15494 Mismatches: 2, R15480C*, T15489A* | - | Genome position: 29263-29282 Mismatches: 1, C29277G* | - | - | - | - |
| USFQ-581 | EPI_ISL_803098 | Genome position:28881-28902 Mismatches: 3,G28881A, G28882A, G28883C | | | | | Genome position: 15469-15494 Mismatches: 2, R15480C*, T15489A* | | Genome position: 29263-29282 Mismatches: 1, C29277G* | | | | |
| USFQ-572 | EPI_ISL_824284 | Genome position:28881-28902 Mismatches: 3,G28881A, G28882A, G28883C | | | | | Genome position: 15469-15494 Mismatches: 2, R15480C*, T15489A* | | Genome position: 29263-29282 Mismatches: 1, C29277G* | | | | |
| USFQ-577 | EPI_ISL_824285 | Genome position:28881-28902 Mismatches: 3,G28881A, G28882A, G28883C | | | | | Genome position: 15469-15494 Mismatches: 2, R15480C*, T15489A* | | Genome position: 29263-29282 Mismatches: 1, C29277G* | | | | |
| USFQ-563 | EPI_ISL_824286 | - | | | | | Genome position: 15469-15494 Mismatches: 2, R15480C*, T15489A* | | Genome position: 29263-29282 Mismatches: 1, C29277G* | | | | |
| USFQ-568 | EPI_ISL_824287 | "Genome position:28881-28902 Mismatches: 3,G28881A, G28882A, G28883C" | | | | | Genome position: 15469-15494 Mismatches: 2, R15480C*, T15489A* | | Genome position: 29263-29282 Mismatches: 1, C29277G* | | | | |
| USFQ-570 | EPI_ISL_824288 | "Genome position:28881-28902 Mismatches: 3,G28881A, G28882A, G28883C" | | | | | Genome position: 15469-15494 Mismatches: 2, R15480C*, T15489A* | | Genome position: 29263-29282 Mismatches: 1, C29277G* | | | | |
| USFQ-571 | EPI_ISL_824289 | "Genome position:28881-28902 Mismatches: 3,G28881A, G28882A, G28883C" | | | | | Genome position: 15469-15494 Mismatches: 2, R15480C*, T15489A* | | Genome position: 29263-29282 Mismatches: 1, C29277G* | | | | |
| USFQ-561 | EPI_ISL_824290 | "Genome position:28881-28902 Mismatches: 3,G28881A, G28882A, G28883C" | | | | | Genome position: 15469-15494 Mismatches: 2, R15480C*, T15489A* | | Genome position: 29263-29282 Mismatches: 1, C29277G* | | | | |
| USFQ-564 | EPI_ISL_824291 | "Genome position:28881-28902 Mismatches: 3,G28881A, G28882A, G28883C" | | | | | Genome position: 15469-15494 Mismatches: 2, R15480C*, T15489A* | | Genome position: 29263-29282 Mismatches: 1, C29277G* | | | | |
| USFQ-569 | EPI_ISL_824292 | "Genome position:28881-28902 Mismatches: 3,G28881A, G28882A, G28883C" | | | | | Genome position: 15469-15494 Mismatches: 2, R15480C*, T15489A* | | Genome position: 29263-29282 Mismatches: 1, C29277G* | | | | |
| USFQ-578 | EPI_ISL_824293 | "Genome position:28881-28902 Mismatches: 3,G28881A, G28882A, G28883C" | | | | | Genome position: 15469-15494 Mismatches: 2, R15480C*, T15489A* | | Genome position: 29263-29282 Mismatches: 1, C29277G* | | | | |
| USFQ-582 | EPI_ISL_824294 | "Genome position:28881-28902 Mismatches: 3,G28881A, G28882A, G28883C" | | | | | Genome position: 15469-15494 Mismatches: 2, R15480C*, T15489A* | | Genome position: 29263-29282 Mismatches: 1, C29277G* | | | | |
