## Supplemental Table 1 for "SARS-CoV-2 genome sequencing from COVID-19 in Ecuadorian patients: a whole country analysis"

**Table1. Mutations found in four SARS CoV2 Genomes from Ecuador compared to Wuhan-Hu-1 (GenBank accession number MN908947)**

| Genome Sequence | | Locality | Clade* | Lineage** | Gen | Mutation | Aminoacid replacement | Type |
|---|---|---|---|---|---|---|---|---|
| ID | GISAID ID | | | | | | | |
| HEE-01 | EPI_ISL_417482 | Quito – Pichincha | 19A | B | nsp1/ORF1ab | T514C | - | - |
| | | | | | S | A25182T | E1207V | Transversions |
| | | | | | nsp3/ORF1ab | C3037T | - | - |
| | | | | | nsp3/ORF1ab | C4010T | L431F | Transition |
| | | | | | nsp12/ORF1ab | C14408T | P323L | Transition |
| | | | | | nsp14/ORF1ab | C18555T | - | - |
| HGSQ-USFQ-018 | EPI_ISL_422563 | Quito – Pichincha | 20B | B.1.1.119 | S | A23403G | D614G | Transition |
| | | | | | ORF3a | A25983G | - | - |
| | | | | | N | G28881A | R203K | Transition |
| | | | | | N | G28882A | R203K | Transition |
| | | | | | N | G28883C | G204R | Transversions |
| | | | | | nsp15/ORF1ab | A1996C | - | - |
| | | | | | nsp15/ORF1ab | C1997T | - | - |
| | | | | | nsp6/ORF1ab | T11737G | N255K | Transversions |
| | | | | | nsp15/ORF1ab | T20497A | S293T | Transversions |
| | | | | | S | G22424T | A288S | Transversions |
| | | | | | S | C24418T | - | - |
| | | | | | ORF3a | A25505C | Q38P | Transversions |
| | | | | | ORF3a | G25647T | L85F | Transversions |
| | | | | | ORF3a | T25676C | L95S | Transition |
| | | | | | ORF3a | C25678T | L96F | Transition |
| | | | | | ORF3a | C25692T | - | - |
| | | | | | ORF3a | T25695G | - | - |
| HGSQ-USFQ-007 | EPI_ISL_422564 | Quito – Pichincha | 19A | B.1.14 | ORF3a | C25708T | L106F | Transition |
| HGSQ-USFQ-010 | EPI_ISL_422565 | Quito – Pichincha | 19A | B.1.14 | ORF3a | C25710T | - | - |
| | | | | | ORF3a | T25711C | Y107H | Transition |
| | | | | | ORF3a | C25728T | - | - |
| | | | | | ORF3a | A25741G | S117G | Transition |
| | | | | | ORF3a | A25756G | R122E | Transition |
| | | | | | ORF3a | G25757A | - | - |
| | | | | | ORF3a | A25764T | - | - |
| | | | | | ORF3a | T25779G | - | - |
| | | | | | ORF3a | G25781A | C130* | Transition |
| | | | | | ORF3a | C25782A | - | - |
| | | | | | ORF3a | G25879A | V163T | Transition |
| | | | | | ORF3a | T25880C | - | - |
| | | | | | ORF3a | C25883A | T164N | Transversions |
| | | | | | ORF3a | T25885G | S165A | Transversions |
| | | | | | ORF8 | A28035G | R48G | Transition |
| | | | | | ORF8 | A28037G | - | - |
| | | | | | ORF8 | G28038T | V49L | Transversions |
| | | | | | ORF8 | C28076T | - | - |
| | | | | | ORF8 | T28078C | V62A | Transition |
| | | | | | nsp3/ORF1ab | C3037T | - | - |
| | | | | | nsp4/ORF1ab | T9172C | - | - |
| | | | | | nsp5/ORF1a | A10948G | - | - |
| | | | | | nsp12/ORF1ab | C14408T | P323L | Transition |
| USFQ-045 | EPI_ISL_471268 | Babahoyo – Los Ríos | 20B | B.1.1.207 | S | A23403G | D614G | Transition |
| | | | | | M | G26730T | V70F | Transversions |
| | | | | | N | G28881A | R203K | Transition |
| | | | | | N | G28882A | R203K | Transition |
| | | | | | N | G28883C | G204R | Transversions |
| | | | | | nsp3/ORF1ab | C3037T | - | - |
| | | | | | nsp12/ORF1ab | C14408T | P323L | Transition |
| USFQ-065 | EPI_ISL_481247 | Babahoyo – Los Ríos | 20B | B.1.1.119 | S | A23403G | D614G | Transition |
| | | | | | N | G28881A | R203K | Transition |
| | | | | | N | G28882A | - | - |
| | | | | | N | G28883C | G204R | Transversions |
| | | | | | nsp3/ORF1ab | C3037T | - | - |
| | | | | | nsp3/ORF1ab | C8266T | - | - |
| | | | | | nsp12/ORF1ab | C14408T | P323L | Transition |
| | | | | | nsp12/ORF1ab | C15222T | - | - |
| USFQ-147 | EPI_ISL_491937 | Calceta – Manabí | 20B | B.1.1.119 | S | A23403G | D614G | Transition |
| | | | | | N | G28881A | R203K | Transition |
| | | | | | N | G28882A | R203K | Transition |
| | | | | | N | G28883C | G204R | Transversions |
| | | | | | nsp3/ORF1ab | C3037T | - | - |
| USFQ-131 | EPI_ISL_491935 | General Villamil Playas – Guayas | 20D | B.1.1.1 | nsp3/ ORF1a | C4002T | T428I | Transition |
| USFQ-194 | EPI_ISL_527810 | Guaranda - Bolívar | 20D | B.1.1.1 | nsp5/ORF1a | G10097A | G15S | Transition |
| USFQ-195 | EPI_ISL_527811 | Guaranda - Bolívar | 20D | B.1.1.1 | nsp12/ORF1a | C13536T | - | - |
| USFQ-212 | EPI_ISL_539785 | Cuenca - Azuay | 20D | B.1.1.1 | nsp12/ORF1ab | C14408T | P323L | Transition |
| USFQ-219 | EPI_ISL_539789 | Macara - Loja | 20D | B.1.1.1 | S | A23403G | D614G | Transition |
| USFQ-513 | EPI_ISL_660537 | Machala - El Oro | 20D | B.1.1.1 | S | C23731T | - | - |
| | | | | | N | G28881A | R203K | Transition |
| | | | | | N | G28882A | R203K | Transition |
| | | | | | N | G28883C | G204R | Transversions |
| | | | | | nsp3/ORF1ab | C3037T | - | - |
| USFQ-105 | EPI_ISL_486842 | Riobamba – Chimborazo | 20B | B.1.1.207 | nsp4/ORF1ab | T9172C | - | - |
| USFQ-108 | EPI_ISL_486843 | Riobamba – Chimborazo | 20B | B.1.1.207 | nsp5/ORF1a | A10948G | - | - |
| USFQ-109 | EPI_ISL_486844 | Riobamba – Chimborazo | 20B | B.1.1.207 | nsp12/ORF1ab | C14408T | P323L | Transition |
| USFQ-111 | EPI_ISL_486846 | Ibarra – Imbabura | 20B | B.1.1.207 | S | A23403G | D614G | Transition |
| USFQ-114 | EPI_ISL_486849 | Ibarra – Imbabura | 20B | B.1.1.207 | M | G26730T | V70F | Transversions |
| USFQ-118 | EPI_ISL_486850 | Ibarra – Imbabura | 20B | B.1.1.207 | N | G28881A | R203K | Transition |
| USFQ-119 | EPI_ISL_486851 | Urcuqui – Imbabura | 20B | B.1.1.207 | N | G28882A | R203K | Transition |
| | | | | | N | G28883C | G204R | Transversions |
| | | | | | nsp3/ORF1ab | C3037T | - | - |
| | | | | | nsp12/ORF1ab | C14408T | P323L | Transition |
| | | | | | nsp14/ORF1ab | C19170T | - | - |
| USFQ-129 | EPI_ISL_491934 | Guayaquil – Guayas | 20B | B.1.1.10 | nsp14/ORF1ab | G19509A | - | - |
| | | | | | S | A23403G | D614G | Transition |
| | | | | | N | G28881A | R203K | Transition |
| | | | | | N | G28882A | R203K | Transition |
| | | | | | N | G28883C | G204R | Transversions |
| | | | | | nsp3/ORF1ab | C3037T | - | - |
| | | | | | nsp12/ORF1a | C14408T | P323L | Transition |
| | | | | | nsp16 /ORF1a | C21242T | T195I | Transition |
| USFQ-112 | EPI_ISL_471270 | Quito – Pichincha | 20B | B.1.1.119 | S | A23403G | D614G | Transition |
| USFQ-106 | EPI_ISL_471271 | Quito – Pichincha | 20B | B.1.1.119 | ORF3a | G25947T | Q185H | Transversions |
| USFQ-707 | EPI_ISL_471269 | Quito – Pichincha | 20B | B.1.1.119 | N | G28881A | R203K | Transition |
| | | | | | N | G28882A | R203K | Transition |
| | | | | | N | G28883C | G204R | Transversions |
| | | | | | nsp3/ORF1ab | C3037T | - | - |
| | | | | | nsp3/ORF1ab | T3788C | - | - |
| | | | | | nsp3/ ORF1a | G5561A | E948K | Transition |
| USFQ-054 | EPI_ISL_481246 | Babahoyo – Los Ríos | 20B | B.1.1.119 | nsp12/ORF1ab | C14408T | P323L | Transition |
| USFQ-066 | EPI_ISL_481248 | Babahoyo – Los Ríos | 20B | B.1.1.119 | nsp16 /ORF1a | C21242T | T195I | Transition |
| | | | | | S | A23403G | D614G | Transition |
| | | | | | N | G28881A | R203K | Transition |
| | | | | | N | G28882A | R203K | Transition |
| | | | | | N | G28883C | G204R | Transversions |
| | | | | | nsp2/ORF1ab | G922A | - | - |
| | | | | | nsp3/ORF1ab | C3037T | - | - |
| | | | | | nsp12/ORF1ab | C14408T | P323L | Transition |
| | | | | | nsp14/ORF1ab | T18436C | F133L | Transition |
| USFQ-120 | EPI_ISL_491932 | Awa indigenous community - Carchi | 20B | B.1.1.119 | nsp16 /ORF1a | C21242T | T195I | Transition |
| | | | | | S | A23403G | D614G | Transition |
| | | | | | N | G28881A | R203K | Transition |
| | | | | | N | G28882A | R203K | Transition |
| | | | | | N | G28883C | G204R | Transversions |
| | | | | | nsp3/ORF1ab | C3037T | - | - |
| | | | | | nsp3/ORF1ab | C3130T | - | - |
| USFQ-026 | EPI_ISL_481244 | Babahoyo – Los Ríos | 20A | B.1 | nsp3/ORF1ab | T3793C | - | - |
| USFQ-113 | EPI_ISL_486848 | Ibarra – Imbabura | 20A | B.1 | nsp12/ORF1ab | C14408T | P323L | Transition |
| | | | | | S | A23403G | D614G | Transition |
| | | | | | M | A26530G | D3G | Transition |
| | | | | | nsp3/ORF1ab | C3037T | - | - |
| USFQ-128 | EPI_ISL_491933 | Guayaquil – Guayas | 20A | B.1 | nsp12/ORF1ab | C14408T | P323L | Transition |
| USFQ-139 | EPI_ISL_491936 | Portoviejo – Manabí | 20A | B.1 | nsp15/ORF1ab | C20148T | - | - |
| USFQ-150 | EPI_ISL_491938 | Manta – Manabí | 20A | B.1 | S | A23403G | D614G | Transition |
| | | | | | M | A26530G | D3G | Transition |
| | | | | | N | C28826T | R185C | Transition |
| | | | | | nsp1/ORF1a | C1059T | T85I | Transition |
| | | | | | nsp3/ORF1ab | C3037T | - | - |
| | | | | | nsp9/ORF1ab | C12880T | - | - |
| USFQ-096 | EPI_ISL_477015 | Quito – Pichincha | 20C | B.1.371 | nsp12/ORF1ab | C14408T | P323L | Transition |

| | | | | | | | | |
|---|---|---|---|---|---|---|---|---|
| | | | | | S | A23403G | D614G | Transition |
| | | | | | ORF3a | G25563T | Q57H | Transversions |
| | | | | | N | G29553A | | |
| | | | | | nsp3/ORF1ab | C3037T | - | - |
| | | | | | nsp5/ORF1a | A10323G | K90R | Transition |
| USFQ-126 | EPI_ISL_491939 | Waorani indigenous community - Orellana | 20A | B.1 | nsp12/ORF1ab | C14408T | P323L | Transition |
| USFQ-127 | EPI_ISL_491940 | Waorani indigenous community - Orellana | 20A | B.1 | nsp15/ORF1ab | A20268G | - | - |
| | | | | | S | A23403G | D614G | Transition |
| | | | | | nsp3/ORF1ab | C3037T | - | - |
| | | | | | nsp3/ ORF1a | C8025T | A1769V | Transition |
| USFQ-097 | EPI_ISL_477016 | Quito – Pichincha | 20A | B.1 | nsp7/ORF1ab | A11983G | - | - |
| | | | | | nsp12/ORF1ab | C14408T | P323L | Transition |
| | | | | | nsp15/ORF1ab | A20268G | - | - |
| | | | | | S | A23403G | D614G | Transition |
| | | | | | nsp3/ORF1ab | C3037T | - | - |
| | | | | | nsp9/ORF1ab | C12915T | - | - |
| USFQ-133 | EPI_ISL_491941 | alapagos Islands - Puerto Baquerizo Morer | 20A | B.1 | nsp12/ORF1ab | C14408T | P323L | Transition |
| | | | | | nsp15/ORF1ab | A20268G | - | - |
| | | | | | S | A23403G | D614G | Transition |
| USFQ-020 | EPI_ISL_471267 | Babahoyo – Los Ríos | 20A | B.1.67 | nsp3/ORF1ab | C3037T | - | - |
| USFQ-004 | EPI_ISL_477014 | Quito – Pichincha | 20A | B.1.67 | nsp12/ORF1ab | C14408T | P323L | Transition |
| USFQ-039 | EPI_ISL_481245 | Babahoyo – Los Ríos | 20A | B.1.67 | S | A23403G | D614G | Transition |
| USFQ-110 | EPI_ISL_486845 | Cayambe – Pichincha | 20A | B.1.67 | M | T26512C | - | - |
| USFQ-1112 | EPI_ISL_486847 | Ibarra – Imbabura | 20A | B.1.67 | | | | |
| | | | | | nsp2/ORF1ab | C1059T | T85I | Transition |
| | | | | | nsp3/ORF1ab | C3037T | - | - |
| USFQ-161 | EPI_ISL_516648 | Yantzaza - Zamora-Chinchipe | 20C | B.1.308 | nsp3/ORF1ab | C6363T | A1215V | Transition |
| | | | | | nsp4/ORF1ab | C9866T | L438F | Transition |
| | | | | | nsp12/ORF1ab | C14408T | P323L | Transition |
| | | | | | nsp12/ORF1ab | A14747G | E436G | Transition |
| | | | | | nsp16/ORF1ab | A20755C | S33R | Transversion |
| | | | | | S | A23403G | D614G | Transition |
| | | | | | ORF3a | C25480T | R30C | Transition |
| | | | | | ORF3a | G25563T | Q57H | Transversion |
| | | | | | ORF7b | A27763G | E3G | Transition |
| | | | | | nsp3/ORF1ab | C3037T | - | - |
| | | | | | nsp3/ORF1ab | A3215G | S166G | Transition |
| USFQ-162 | EPI_ISL_516649 | Yantzaza - Zamora-Chinchipe | 20A | B.1.223 | nsp12/ORF1ab | C14408T | P323L | Transition |
| USFQ-197 | EPI_ISL_527813 | Esmeraldas - Esmeraldas | 20A | B.1.223 | nsp12/ORF1ab | G15438T | M666I | Transversion |
| | | | | | nsp15/ORF1ab | A20268G | - | - |
| | | | | | S | A23403G | D614G | Transition |
| | | | | | nsp2/ORF1ab | C1457T | R218C | Transition |
| | | | | | nsp4/ORF1ab | C8782T | - | - |
| USFQ-165 | EPI_ISL_516650 | Quito-Pichincha | 19B | A.1 | nsp4/ORF1ab | T9445C | - | - |
| | | | | | nsp13/ORF1ab | T17531C | I432T | Transition |
| | | | | | nsp13/ORF1ab | C17747T | P504L | Transition |
| | | | | | nsp13/ORF1ab | A17858G | Y541C | Transition |
| | | | | | nsp14/ORF1ab | C18060T | - | - |
| | | | | | nsp14/ORF1ab | G18756T | - | - |
| | | | | | S | A24694T | - | - |
| | | | | | ORF8 | T28144C | L84S | Transition |
| | | | | | nsp3/ORF1ab | T2987C | F90L | Transition |
| USFQ-167 | EPI_ISL_516651 | Quevedo - Los Ríos | 20A | B.1.223 | nsp3/ORF1ab | C3037T | - | - |
| USFQ-171 | EPI_ISL_516652 | Coca - Orellana | 20A | B.1.223 | nsp3/ORF1ab | A3215G | S166G | Transition |
| USFQ-186 | EPI_ISL_525436 | Otavalo - Imbabura | 20A | B.1.223 | nsp12/ORF1ab | C14408T | P323L | Transition |
| USFQ-MC01 | EPI_ISL_525438 | Quito - Pichincha | 20A | B.1.223 | nsp12/ORF1ab | G15438T | M666I | Transversion |
| USFQ-216 | EPI_ISL_539788 | Cuenca - Azuay | 20A | B.1.223 | nsp15/ORF1ab | A20268G | - | - |
| | | | | | S | A23403G | D614G | Transition |
| | | | | | nsp2/ORF1ab | C2113T | - | - |
| | | | | | nsp3/ORF1ab | C3037T | - | - |
| USFQ-163 | EPI_ISL_525430 | Quito - Pichincha | 20A | B.1.9 | nsp3/ORF1ab | C7765T | - | - |
| | | | | | nsp12/ORF1ab | C14408T | P323L | Transition |
| | | | | | nsp13/ORF1ab | C17690T | S485L | Transition |
| | | | | | nsp14/ORF1ab | C18877T | - | - |
| | | | | | S | A23403G | D614G | Transition |
| | | | | | ORF3a | G25563T | Q57H | Transversion |
| | | | | | nsp3/ORF1ab | C3037T | - | - |
| USFQ-177 | EPI_ISL_525431 | Zamora - Zamora-Chinchipe | 20A | B.1 | nsp3/ORF1ab | C3130T | - | - |
| USFQ-184 | EPI_ISL_525435 | Orellana - Orellana | 20A | B.1 | nsp3/ORF1ab | G3403T | - | - |
| USFQ-188 | EPI_ISL_525437 | Otavalo - Imbabura | 20A | B.1 | nsp12/ORF1ab | C14408T | P323L | Transition |
| | | | | | S | A23403G | D614G | Transition |
| | | | | | M | A26530G | D3G | Transition |
| USFQ-193 | EPI_ISL_527809 | Guaranda - Bolívar | 20B | B.1.1.119 | nsp3/ORF1ab | C3037T | - | - |
| USFQ-201 | EPI_ISL_527815 | Esmeraldas - Esmeraldas | 20B | B.1.1.119 | nsp12/ORF1ab | T13552C | Y38H | Transition |
| USFQ-203 | EPI_ISL_527816 | Esmeraldas - Esmeraldas | 20B | B.1.1.119 | nsp12/ORF1ab | C14408T | P323L | Transition |
| USFQ-207 | EPI_ISL_527819 | Guayaquil – Guayas | 20B | B.1.1.119 | nsp16/ORF1ab | C21242T | T195I | Transition |
| USFQ-249 | EPI_ISL_697787 | Azogues - Cañar | 20B | B.1.1.119 | S | A23403G | D614G | Transition |
| USFQ-528 | EPI_ISL_697797 | Guayaquil - Guayas | 20B | B.1.1.119 | N | G28881A | R203K | Transition |
| USFQ-228 | EPI_ISL_660529 | Ambato - Tungurahua | 20B | B.1.1.119 | N | G28882A | R203K | Transition |
| | | | | | N | G28883C | G204R | Transversion |
| USFQ-196 | EPI_ISL_527812 | Esmeraldas - Esmeraldas | 20A | B.1 | nsp3/ORF1ab | C3037T | - | - |
| USFQ-199 | EPI_ISL_527814 | Esmeraldas - Esmeraldas | 20A | B.1 | nsp3/ORF1ab | C3130T | - | - |
| USFQ-204 | EPI_ISL_527817 | Esmeraldas - Esmeraldas | 20A | B.1 | nsp3/ORF1ab | T3793C | - | - |
| USFQ-214 | EPI_ISL_539787 | Cuenca - Azuay | 20A | B.1 | nsp12/ORF1ab | C14408T | P323L | Transition |
| USFQ-222 | EPI_ISL_539791 | Macara - Loja | 20A | B.1 | S | A23403G | D614G | Transition |
| USFQ-224 | EPI_ISL_539792 | Catamayo - Loja | 20A | B.1 | M | A26530G | D3G | Transition |
| USFQ-402 | EPI_ISL_660532 | Puyo - Pastaza | 20A | B.1 | | | | |
| | | | | | nsp2/ORF1ab | C1059T | T85I | Transition |
| | | | | | nsp3/ORF1ab | C3037T | - | - |
| USFQ-178 | EPI_ISL_525432 | El Pangui - Zamora-Chinchipe | 20C | B.1 | nsp12/ORF1ab | C14408T | P323L | Transition |
| USFQ-182 | EPI_ISL_525433 | Quevedo - Los Ríos | 20C | B.1 | nsp12/ORF1ab | A14747G | E436G | Transition |
| | | | | | nsp13/ORF1ab | C16658T | T141I | Transition |
| | | | | | nsp16/ORF1ab | A20755C | S33R | Transversion |
| | | | | | S | A23403G | D614G | Transition |
| | | | | | ORF3a | G25563T | Q57H | Transversion |
| | | | | | nsp3/ORF1ab | C3037T | - | - |
| | | | | | nsp3/ORF1ab | C4002T | T428I | Transition |
| USFQ-183 | EPI_ISL_525434 | Quevedo - Los Ríos | 20D | B.1.1.1 | nsp3/ORF1ab | T4092C | I458T | Transition |
| | | | | | nsp5/ORF1ab | G10097A | G15S | Transition |
| | | | | | nsp12/ORF1ab | C13536T | - | - |
| | | | | | nsp12/ORF1ab | C14408T | P323L | Transition |
| | | | | | S | A23403G | D614G | Transition |
| | | | | | S | C23731T | - | - |
| | | | | | N | G28881A | R203K | Transition |
| | | | | | N | G28882A | R203K | Transition |
| | | | | | N | G28883C | G204R | Transversion |
| | | | | | nsp3/ORF1ab | C3037T | - | - |
| | | | | | nsp3/ORF1ab | C3040T | - | - |
| USFQ-206 | EPI_ISL_527818 | Guayaquil - Guayas | 20B | B.1.1.119 | nsp7/ORF1ab | A11983G | - | - |
| | | | | | nsp12/ORF1ab | T13552C | Y38H | Transition |
| | | | | | nsp12/ORF1ab | C14408T | P323L | Transition |
| | | | | | nsp16/ORF1ab | C21242T | T195I | Transition |
| | | | | | S | A23403G | D614G | Transition |
| | | | | | N | G28881A | R203K | Transition |
| | | | | | N | G28882A | R203K | Transition |
| | | | | | N | G28883C | G204R | Transversion |
| | | | | | nsp3/ORF1ab | C3037T | - | - |
| | | | | | nsp12/ORF1ab | T14079C | - | - |
| USFQ-208 | EPI_ISL_539783 | Tena - Napo | 20B | B.1.1.119 | nsp12/ORF1ab | C14408T | P323L | Transition |
| USFQ-527 | EPI_ISL_697796 | Tena - Napo | 20B | B.1.1.119 | S | A23403G | D614G | Transition |
| | | | | | ORF3a | G25690T | G100C | Transversion |
| | | | | | N | G28881A | R203K | Transition |
| | | | | | N | G28882A | R203K | Transition |
| | | | | | N | G28883C | G204R | Transversion |
| | | | | | nsp3/ORF1ab | C3037T | - | - |
| USFQ-209 | EPI_ISL_539784 | Tena - Napo | 20A | B.1 | nsp5/ORF1ab | A10323G | K90R | Transition |
| USFQ-213 | EPI_ISL_539786 | Cuenca - Azuay | 20A | B.1 | nsp12/ORF1ab | C14408T | P323L | Transition |
| USFQ-220 | EPI_ISL_539790 | Macara - Loja | 20A | B.1 | nsp15/ORF1ab | A20268G | - | - |
| | | | | | S | A23403G | D614G | Transition |
| | | | | | nsp2ORF1ab | G922A | - | - |
| USFQ-226 | EPI_ISL_539793 | Catamayo - Loja | 20B | B.1.1.119 | nsp3/ORF1ab | C3037T | - | - |
| USFQ-253 | EPI_ISL_574431 | Babahoyo - Los Ríos | 20B | B.1.1.119 | nsp12/ORF1ab | C14408T | P323L | Transition |
| | | | | | nsp14/ORF1ab | T18436C | F133L | Transition |
| | | | | | nsp16/ORF1ab | C21242T | T195I | Transition |
| | | | | | S | A23403G | D614G | Transition |
| | | | | | N | G28881A | R203K | Transition |
| | | | | | N | G28882A | R203K | Transition |
| | | | | | N | G28883C | G204R | Transversion |
| USFQ-173 | EPI_ISL_697783 | Ibarra-Imbabura | 20B | B.1.1.119 | nsp1/ORF1ab | C292T | - | - |
| USFQ-346 | EPI_ISL_697790 | Otavalo - Imbabura | 20B | B.1.1.119 | nsp3/ORF1ab | C3037T | - | - |

| | | | | | | | | |
|---|---|---|---|---|---|---|---|---|
| USFQ-405 | EPI_ISL_697792 | San Pablo - Imbabura | 20B | B.1.1.119 | nsp3/ORF1ab | G4319A | A534T | Transition |
| | | | | | nsp3/ORF1ab | C5842T | - | - |
| | | | | | nsp12/ORF1ab | T13935C | - | - |
| | | | | | nsp12/ORF1ab | C14408T | P323L | Transition |
| | | | | | nsp13/ORF1ab | C16750T | P172S | Transition |
| | | | | | nsp14/ORF1ab | G19509A | - | - |
| | | | | | S | A23403G | D614G | Transition |
| | | | | | S | C24034T | - | - |
| | | | | | ORF8 | T28009C | I39T | Transition |
| | | | | | N | G28881A | R203K | Transition |
| | | | | | N | G28882A | R203K | Transition |
| | | | | | N | G28883C | G204R | Transversion |
| USFQ-185 | EPI_ISL_697784 | Otavalo - Imbabura | 20D | B.1.1.1 | nsp3/ORF1ab | C3037T | - | - |
| USFQ-189 | EPI_ISL_697786 | Otavalo - Imbabura | 20D | B.1.1.1 | nsp3/ORF1ab | A3237G | Q173R | Transition |
| | | | | | nsp3/ORF1ab | C4002T | T428I | Transition |
| | | | | | nsp3/ORF1ab | C4878T | T720I | Transition |
| | | | | | nsp3/ORF1ab | G5515T | - | - |
| | | | | | nsp5/ORF1ab | C10078T | - | - |
| | | | | | nsp5/ORF1ab | G10097A | G15S | Transition |
| | | | | | nsp6/ORF1ab | C11758T | - | - |
| | | | | | nsp6/ORF1ab | C11812T | - | - |
| | | | | | nsp12/ORF1ab | C13536T | - | - |
| | | | | | nsp12/ORF1ab | C14408T | P323L | Transition |
| | | | | | nsp15/ORF1ab | G20434T | D272Y | Transversion |
| | | | | | S | A23403G | D614G | Transition |
| | | | | | S | C23731T | - | - |
| | | | | | N | G28881A | R203K | Transition |
| | | | | | N | G28882A | R203K | Transition |
| | | | | | N | G28883C | G204R | Transversion |
| USFQ-344 | EPI_ISL_697789 | Otavalo - Imbabura | 20A | B.1.6 | nsp3/ORF1ab | C3037T | - | - |
| | | | | | nsp3/ORF1ab | C5974T | - | - |
| | | | | | nsp3/ORF1ab | C8394T | A1892V | |
| | | | | | nsp8/ORF1ab | C12469T | - | - |
| | | | | | nsp12/ORF1ab | C14408T | P323L | Transition |
| | | | | | nsp12/ORF1ab | C15277T | H613Y | Transition |
| | | | | | nsp12/ORF1ab | C15324T | - | - |
| | | | | | S | A23403G | D614G | Transition |
| USFQ-381 | EPI_ISL_697791 | Otavalo - Imbabura | 20B | B.1.1.119 | nsp2/ORF1ab | A2653G | - | - |
| | | | | | nsp3/ORF1ab | C3037T | - | - |
| | | | | | nsp3/ORF1ab | C5986T | - | - |
| | | | | | nsp4/ORF1ab | T9952C | - | - |
| | | | | | nsp7/ORF1ab | A11983G | - | - |
| | | | | | nsp12/ORF1ab | T14103C | - | - |
| | | | | | nsp12/ORF1ab | C14408T | P323L | Transition |
| | | | | | nsp15/ORF1ab | A19812C | K64N | Transversion |
| | | | | | S | C21614T | L18F | Transition |
| | | | | | S | A23403G | D614G | Transition |
| | | | | | S | A25122G | N1187S | Transition |
| | | | | | ORF3a | G25593T | K67N | Transversion |
| | | | | | ORF7a | C27476T | T28I | Transition |
| | | | | | N | G28881A | R203K | Transition |
| | | | | | N | G28882A | R203K | Transition |
| | | | | | N | G28883C | G204R | Transversion |
| USFQ-433 | EPI_ISL_697793 | Otavalo - Imbabura | 20B | B.1.1.119 | nsp2/ORF1a | G922A | - | - |
| | | | | | nsp3/ORF1ab | C3037T | - | - |
| | | | | | nsp3/ORF1ab | A5209C | - | - |
| | | | | | nsp3/ORF1ab | A6613G | - | - |
| | | | | | nsp4/ORF1ab | G9928T | M458I | Transversion |
| | | | | | nsp12/ORF1ab | C14408T | P323L | Transition |
| | | | | | nsp13/ORF1ab | T17475C | - | - |
| | | | | | nsp14/ORF1ab | T18436C | F133L | Transition |
| | | | | | nsp16/ORF1ab | C21242T | T195I | Transition |
| | | | | | S | A23403G | D614G | Transition |
| | | | | | N | G28881A | R203K | Transition |
| | | | | | N | G28882A | R203K | Transition |
| | | | | | N | G28883C | G204R | Transversion |
| USFQ-523 | EPI_ISL_697794 | Morona – Morona Santiago | 20D | B.1.1.1 | nsp3/ORF1ab | C3037T | - | - |
| | | | | | nsp3/ORF1ab | C4002T | T428I | Transition |
| | | | | | nsp5/ORF1ab | G10097A | G15S | Transition |
| | | | | | nsp5/ORF1ab | T10459C | - | - |
| | | | | | nsp12/ORF1ab | C13536T | - | - |
| | | | | | nsp12/ORF1ab | C14408T | P323L | Transition |
| | | | | | S | A23403G | D614G | Transition |
| | | | | | S | C23731T | - | - |
| | | | | | N | G28881A | R203K | Transition |
| | | | | | N | G28882A | R203K | Transition |
| | | | | | N | G28883C | G204R | Transversion |
| USFQ-524 | EPI_ISL_697795 | Morona – Morona Santiago | 20B | B.1.1.119 | nsp2/ORF1a | G922A | - | - |
| | | | | | nsp3/ORF1ab | C3037T | - | - |
| | | | | | nsp5/ORF1ab | T10480C | - | - |
| | | | | | nsp8/ORF1ab | A12358T | - | - |
| | | | | | nsp12/ORF1ab | G13812T | M124I | Transversion |
| | | | | | nsp12/ORF1ab | C14408T | P323L | Transition |
| | | | | | nsp12/ORF1ab | C15352T | L638F | Transition |
| | | | | | nsp13/ORF1ab | C18032T | T599I | Transition |
| | | | | | nsp14/ORF1ab | T18436C | F133L | Transition |
| | | | | | nsp16/ORF1ab | C21242T | T195I | Transition |
| | | | | | S | A23403G | D614G | Transition |
| | | | | | N | G28881A | R203K | Transition |
| | | | | | N | G28882A | R203K | Transition |
| | | | | | N | G28883C | G204R | Transversion |
| USFQ-1188 | EPI_ISL_697785 | Otavalo - Imbabura | 20B | B.1.1.119 | nsp3/ORF1ab | C3037T | - | - |
| USFQ-322 | EPI_ISL_697788 | Otavalo - Imbabura | 20B | B.1.1.119 | nsp3/ORF1ab | C4276T | - | - |
| | | | | | nsp12/ORF1ab | C14408T | P323L | Transition |
| | | | | | nsp16/ORF1ab | C21242T | T195I | Transition |
| | | | | | S | A23403G | D614G | Transition |
| | | | | | N | G28881A | R203K | Transition |
| | | | | | N | G28882A | R203K | Transition |
| | | | | | N | G28883C | G204R | Transversion |
| USFQ-531 | EPI_ISL_697798 | Sucumbios | 20A | B.1 | nsp3/ORF1ab | C3037T | - | - |
| USFQ-533 | EPI_ISL_697799 | Sucumbios | 20A | B.1 | nsp3/ORF1ab | T3793C | - | - |
| USFQ-539 | EPI_ISL_697800 | Santa Elena | 20A | B.1 | nsp12/ORF1ab | C14408T | P323L | Transition |
| | | | | | nsp13/ORF1ab | C18032T | T599I | Transition |
| | | | | | S | A23403G | D614G | Transition |
| | | | | | M | A26530G | D3G | Transition |
| USFQ-553 | EPI_ISL_728202 | Santo Domingo – Santo Domingo | 20B | B.1.1.159 | nsp2/ORF1ab | C1191T | P129L | Transition |
| USFQ-555 | EPI_ISL_728203 | Santo Domingo – Santo Domingo | 20B | B.1.1.159 | nsp3/ORF1ab | C3037T | - | - |
| USFQ-558 | EPI_ISL_728205 | Sucumbios | 20B | B.1.1.159 | nsp3/ORF1ab | C7113T | T1465I | Transition |
| | | | | | nsp5/ORF1ab | G10360A | - | - |
| | | | | | nsp6/ORF1ab | G11540T | V190F | Transversion |
| | | | | | nsp12/ORF1ab | C14408T | P323L | Transition |
| | | | | | nsp14/ORF1ab | C18131T | T31I | Transition |
| | | | | | nsp16/ORF1ab | G21538T | V294F | Transversion |
| | | | | | S | A23403G | D614G | Transition |
| | | | | | N | C28290T | P6L | Transition |
| | | | | | N | G28881A | R203K | Transition |
| | | | | | N | G28882A | R203K | Transition |
| | | | | | N | G28883C | G204R | Transversion |
| USFQ-556 | EPI_ISL_728204 | Santo Domingo – Santo Domingo | 19B | A | nsp1/ORF1ab | C275T | L4F | Transition |
| | | | | | nsp1/ORF1ab | T490A | D75E | Transversion |
| | | | | | nsp2/ORF1ab | C1567T | - | - |
| | | | | | nsp3/ORF1ab | C3165T | A149V | Transition |
| | | | | | nsp3/ORF1ab | C4543T | - | - |
| | | | | | nsp3/ORF1ab | A5234G | K839E | Transition |
| | | | | | nsp4/ORF1ab | C8782T | - | - |
| | | | | | nsp4/ORF1ab | T9477A | F308Y | Transversion |
| | | | | | nsp4/ORF1ab | C10029T | T492I | Transition |
| | | | | | nsp6/ORF1ab | C11005A | H11Q | Transversion |
| | | | | | nsp12/ORF1ab | C14805T | - | - |
| | | | | | nsp12/ORF1ab | C15960T | - | - |
| | | | | | nsp13/ORF1ab | C16466T | P77L | Transition |
| | | | | | nsp14/ORF1ab | C18115T | H26Y | Transition |
| | | | | | nsp14/ORF1ab | T18417C | - | - |
| | | | | | nsp14/ORF1ab | T18736C | F233L | Transition |
| | | | | | S | A22525G | - | - |
| | | | | | S | C24034T | - | - |
| | | | | | S | G24697A | - | - |
| | | | | | ORF8 | G28077C | V62L | Transversion |
| | | | | | ORF8 | T28144C | L84S | Transition |
| | | | | | N | A28272T | - | - |

| | | | | | | | | |
|---|---|---|---|---|---|---|---|---|
| | | | | | N | C28657T | - | - |
| | | | | | N | G28975T | M234I | Transversion |
| USFQ-520 | EPI_ISL_660539 | Puyo - Pastaza | 20B | B.1.1.119 | nsp3/ORF1ab | C3037T | - | - |
| | | | | | nsp6/ORF1ab | A11326G | - | - |
| | | | | | nsp9/ORF1ab | G12769T | - | - |
| | | | | | nsp12/ORF1ab | C13862T | T141I | Transition |
| | | | | | nsp12/ORF1ab | C14408T | P323L | Transition |
| | | | | | nsp14/ORF1ab | G19509A | - | - |
| | | | | | nsp15/ORF1ab | A20539G | K307E | Transition |
| | | | | | S | A23403G | D614G | Transition |
| | | | | | N | G28881A | R203K | Transition |
| | | | | | N | G28882A | R203K | Transition |
| | | | | | N | G28883C | G204R | Transversion |
| USFQ-515 | EPI_ISL_660538 | Machala - El Oro | 20A | B.1 | nsp3/ORF1ab | C3037T | - | - |
| USFQ-509 | EPI_ISL_660536 | Machala - El Oro | 20A | B.1 | nsp3/ORF1ab | T3793C | - | - |
| USFQ-487 | EPI_ISL_660534 | Latacunga - Cotopaxi | 20A | B.1 | nsp7/ORF1ab | A11983G | - | - |
| | | | | | nsp12/ORF1ab | C14408T | P323L | Transition |
| | | | | | nsp12/ORF1ab | G15957A | - | - |
| | | | | | nsp14/ORF1ab | C18928T | P297S | Transition |
| | | | | | S | A23403G | D614G | Transition |
| | | | | | M | A26530G | D3G | Transition |
| USFQ-506 | EPI_ISL_660535 | Machala - El Oro | 20A | B.1 | nsp3/ORF1ab | C3037T | - | - |
| | | | | | nsp3/ORF1ab | C7101T | A1461V | Transition |
| | | | | | nsp3/ORF1ab | G7798T | K1693N | Transversion |
| | | | | | nsp3/ ORF1a | C8025T | A1769V | Transition |
| | | | | | nsp12/ORF1ab | C14408T | P323L | Transition |
| | | | | | nsp15/ORF1ab | A20268G | - | - |
| | | | | | S | A23403G | D614G | Transition |
| | | | | | ORF8 | C28201T | S103L | Transition |
| | | | | | N | C28409T | P46S | Transition |
| USFQ-485 | EPI_ISL_660533 | Latacunga - Cotopaxi | 20A | B.1.197 | nsp2/ORF1ab | A2442G | E546G | Transition |
| | | | | | nsp3/ORF1ab | C3037T | - | - |
| | | | | | nsp3/ORF1ab | C6723T | T1335I | Transition |
| | | | | | nsp12/ORF1ab | C14408T | P323L | Transition |
| | | | | | nsp12/ORF1ab | T14895C | - | - |
| | | | | | nsp13/ORF1ab | C17421T | - | - |
| | | | | | S | A23403G | D614G | Transition |
| | | | | | ORF3a | T26094C | - | - |
| | | | | | ORF6 | T27222C | - | - |
| USFQ-231 | EPI_ISL_660531 | Ambato - Tungurahua | 20A | B.1.197 | nsp3/ORF1ab | C3037T | - | - |
| | | | | | nsp3/ORF1ab | C6723T | T1335I | Transition |
| | | | | | nsp12/ORF1ab | C14408T | P323L | Transition |
| | | | | | nsp12/ORF1ab | T14895C | - | - |
| | | | | | nsp13/ORF1ab | C17421T | - | - |
| | | | | | S | A23403G | D614G | Transition |
| | | | | | ORF3a | T26094C | - | - |
| | | | | | ORF6 | T27222C | - | - |
| USFQ-230 | EPI_ISL_660530 | Ambato - Tungurahua | 20B | B.1.1.10 | nsp2/ORF1ab | A2653G | - | - |
| | | | | | nsp3/ORF1ab | C3037T | - | - |
| | | | | | nsp12/ORF1ab | C14408T | P323L | Transition |
| | | | | | nsp14/ORF1ab | C19170T | - | - |
| | | | | | nsp14/ORF1ab | G19509A | - | - |
| | | | | | S | A23403G | D614G | Transition |
| | | | | | N | G28881A | R203K | Transition |
| | | | | | N | G28882A | R203K | Transition |
| | | | | | N | G28883C | G204R | Transversion |
| USFQ-581 | EPI_ISL_803098 | Valencia - Los Ríos | 20I/501Y.V1 | B.1.1.7 | nsp2/ORF1ab | C913T | - | - |
| USFQ-578 | EPI_ISL_824293 | Valencia - Los Ríos | 20I/501Y.V1 | B.1.1.7 | nsp3/ORF1ab | C3037T | - | - |
| USFQ-582 | EPI_ISL_824294 | Valencia - Los Ríos | 20I/501Y.V1 | B.1.1.7 | nsp3/ORF1ab | C3267T | T183I | Transition |
| | | | | | nsp3/ORF1ab | C5388A | A890D | Transversion |
| | | | | | nsp3/ORF1ab | C5986T | - | - |
| | | | | | nsp3/ORF1ab | T6954C | I1412T | Transition |
| | | | | | nsp12/ORF1ab | C14408T | P323L | Transition |
| | | | | | nsp12/ORF1ab | C14676T | - | - |
| | | | | | nsp12/ORF1ab | C15279T | - | - |
| | | | | | nsp12/ORF1ab | T16176C | - | - |
| | | | | | nsp14/ORF1ab | G19518T | L493F | Transversion |
| | | | | | S | 21765-21770 | non-codon-aligned deletion | - |
| | | | | | S | A23063T | N501Y | Transversion |
| | | | | | S | C23220T | T553I | Transition |
| | | | | | S | C23271A | A570D | Transversion |
| | | | | | S | A23403G | D614G | Transition |
| | | | | | S | C23604A | P681H | Transversion |
| | | | | | S | C23709T | T716I | Transition |
| | | | | | S | T24506G | S982A | Transversion |
| | | | | | S | G24914C | D1118H | Transversion |
| | | | | | ORF8 | C27972T | Q27* | Transition |
| | | | | | ORF8 | G28048T | R52I | Transversion |
| | | | | | ORF8 | A28111G | Y73C | Transition |
| | | | | | N | G28280C | D3L | Transversion |
| | | | | | N | A28281T | D3L | Transversion |
| | | | | | N | T28282A | D3L | Transversion |
| | | | | | N | G28881A | R203K | Transition |
| | | | | | N | G28882A | R203K | Transition |
| | | | | | N | G28883C | G204R | Transversion |
| | | | | | N | C28977T | S235F | Transition |
| USFQ-572 | EPI_ISL_824284 | Santo Domingo - Santo Domingo | 20B | B.1.1.31 | nsp3/ORF1ab | C3037T | - | - |
| USFQ-568 | EPI_ISL_824287 | Otavalo - Imbabura | 20B | B.1.1.31 | nsp3/ORF1ab | C3587T | H290Y | Transition |
| USFQ-564 | EPI_ISL_824291 | Otavalo - Imbabura | 20B | B.1.1.31 | nsp3/ORF1ab | C4754T | P679S | Transition |
| USFQ-569 | EPI_ISL_824292 | Otavalo - Imbabura | 20B | B.1.1.31 | nsp3/ORF1ab | C6026T | P1103S | Transition |
| | | | | | nsp3/ORF1ab | T7624C | - | - |
| | | | | | nsp12/ORF1ab | T13552C | Y38H | Transition |
| | | | | | nsp12/ORF1ab | C14408T | P323L | Transition |
| | | | | | nsp13/ORF1ab | C17373T | - | - |
| | | | | | nsp13/ORF1ab | C17518T | L428F | Transition |
| | | | | | nsp16/ORF1ab | C21242T | T195I | Transition |
| | | | | | S | A23403G | D614G | Transition |
| | | | | | ORF3a | G25855C | D155H | Transversion |
| | | | | | N | G28881A | R203K | Transition |
| | | | | | N | G28882A | R203K | Transition |
| | | | | | N | G28883C | G204R | Transversion |
| | | | | | ORF10 | C29642T | Q29* | Transition |
| USFQ-577 | EPI_ISL_824285 | Santo Domingo - Santo Domingo | 20B | B.1.1.10 | nsp3/ORF1ab | C3037T | - | - |
| USFQ-570 | EPI_ISL_824288 | Santo Domingo - Santo Domingo | 20B | B.1.1.10 | nsp3/ORF1ab | C7392T | P1558L | Transition |
| USFQ-571 | EPI_ISL_824289 | Santo Domingo - Santo Domingo | 20B | B.1.1.10 | nsp6/ORF1ab | G11417T | V149F | Transversion |
| USFQ-561 | EPI_ISL_824290 | Otavalo - Imbabura | 20B | B.1.1.10 | nsp12/ORF1ab | C14408T | P323L | Transition |
| | | | | | nsp12/ORF1ab | T15642C | - | - |
| | | | | | nsp14/ORF1ab | C19170T | - | - |
| | | | | | nsp14/ORF1ab | G19509A | - | - |
| | | | | | nsp15/ORF1ab | G19962T | - | - |
| | | | | | nsp16/ORF1ab | G21468T | M270I | Transversion |
| | | | | | S | A23403G | D614G | Transition |
| | | | | | **S** | **G25062C** | **G1167A** | Transversion |
| | | | | | N | C28278A | S2Y | Transversion |
| | | | | | N | G28881A | R203K | Transition |
| | | | | | N | G28882A | R203K | Transition |
| | | | | | N | G28883C | G204R | Transversion |
| | | | | | N | C29370T | T366I | Transition |
| USFQ-563 | EPI_ISL_824286 | Otavalo - Imbabura | 20A | B.1.325 | nsp3/ORF1ab | C3037T | - | - |
| | | | | | **nsp3/ORF1ab** | **C3326G** | **Q203E** | Transversion |
| | | | | | nsp3/ORF1ab | T3793C | - | - |
| | | | | | nsp12/ORF1ab | C14408T | P323L | Transition |
| | | | | | nsp12/ORF1ab | T14565A | - | - |
| | | | | | S | A23403G | D614G | Transition |
| | | | | | M | A26530G | D3G | Transition |
| | | | | | **ORF8** | **C28227A** | **H112N** | Transversion |

*Defined in NextStain (NextClade) on January 25th
** Defined in GISAID on January 27th
